# Surveillance-adjusted syphilis risk mapping across U.S. counties: a Bayesian spatial analysis with external validation against HIV and gonorrhea outcomes

**DOI:** 10.64898/2026.07.09.26357652

**Authors:** Qiancheng Ma, Tianzhen Zhang, Dongzhi Lin

**Author notes:** Corresponding author: Qiancheng Ma.

## Abstract

**Objectives:** To estimate surveillance-adjusted county-level residual syphilis risk, quantify posterior support for elevated risk, and identify the geographic distribution of stably high-risk areas across the contiguous United States and the District of Columbia.

**Methods:** County-year primary and secondary syphilis counts from 3,109 counties during 2010–2022 were analyzed using a Bayesian negative-binomial spatial model with county-level covariates capturing social vulnerability and healthcare and surveillance related structure. Residual spatial risk, posterior exceedance probabilities, and stably high-risk counties were estimated. External validation examined whether county-level residual syphilis risk was associated with HIV and gonorrhea burden.

**Results:** A total of 850 stably high-risk counties were identified. These counties were concentrated in the southeastern United States and along the Gulf Coast, with additional clusters in the north-central region and along the Atlantic and Pacific coasts. The social vulnerability index showed the strongest positive association with reported syphilis rates, followed by primary care physician density. External validation and sensitivity analyses showed that higher county-level residual syphilis risk estimates were positively associated with higher HIV diagnosis rates and gonorrhea rates, indicating that these estimates were not merely model-derived numerical outputs but were meaningfully related to the county-level distribution of sexually transmitted infection risk. These findings indicate that surveillance-adjusted residual spatial risk estimates and posterior exceedance probabilities may provide useful county-level evidence for syphilis control prioritization and resource allocation.

## 1. Introduction

Reported primary and secondary syphilis has resurged sharply in the United States over the past decade, increasing from 13,627 cases in 2010 to 58,373 cases in 2022[1]. This resurgence has not been evenly distributed, but has instead imposed a disproportionate burden across affected populations and geographic areas, reflecting marked heterogeneity in sexual networks, healthcare access, and local prevention and treatment capacity[2, 3].

A central challenge in interpreting county-level syphilis surveillance data is that reported case counts reflect not only underlying transmission, but also the local capacity to detect, diagnose, treat, and report infections. Access to sexual health services is uneven across communities, shaped by factors such as provider availability, insurance coverage, federally qualified health center (FQHC) distribution, and health professional shortage status. These differences may influence whether infections are identified and entered into surveillance systems. As a result, counties with limited healthcare and public health infrastructure may appear to have low reported rates even when residual transmission risk remains substantial (that is, risk remaining after accounting for observable differences in healthcare access and surveillance capacity), whereas better-resourced counties may appear high-burden partly because they detect and report more cases[4].This creates a surveillance gap problem: observed county-level syphilis rates may conflate true epidemiologic risk with differences in screening, healthcare access, and reporting capacity. Therefore, distinguishing surveillance capacity from residual underlying risk is essential for identifying counties where syphilis burden may be underestimated by routine case counts.

Most previous U.S. syphilis studies have examined local, state-level, or population-specific patterns, leaving limited evidence on national county-level variation in primary and secondary syphilis risk.[5] The closest national county-level study by Chang et al. identified hot spot counties for chlamydia, gonorrhea, and primary and secondary syphilis across the 48 contiguous states using hot spot analysis, and then used spatial logistic regression to examine correlates of hot spot membership. Because covariates were introduced only after hot spot detection, the identified hot spots may partly reflect differences in screening, healthcare access, and reporting capacity rather than underlying transmission alone. We therefore used a Bayesian spatiotemporal framework to model county-year primary and secondary (P&S) syphilis counts directly, adjust for surveillance-related covariates, treat suppressed or unavailable counts as missing, and identify stable elevated residual risk using posterior exceedance probabilities.

In this study, we developed a Bayesian spatiotemporal disease-mapping framework to identify surveillance-adjusted residual spatial risk of primary and secondary syphilis across U.S. counties. Rather than mapping reported rates alone, we modelled county-year syphilis counts while accounting for population size, healthcare access, surveillance-related infrastructure, and spatial and temporal dependence. We defined county-level residual spatial risk using posterior residual relative risk after covariate adjustment and assessed posterior support for elevated risk using posterior exceedance probabilities. We further evaluated whether county-level residual syphilis risk corresponded to HIV and gonorrhea burden in external validation analyses, providing external evidence for the public health relevance of the identified high-risk areas.

## 2. Materials and methods

Detailed model specification, prior distributions, Markov chain Monte Carlo (MCMC) settings, and diagnostic procedures are provided in Supporting information S1 Text .

This county-level ecological study examined reported primary and secondary syphilis burden across 3,109 counties in the contiguous United States and the District of Columbia from 2010 to 2022, with county-year as the unit of analysis. **Annual syphilis case counts** were obtained from CDC AtlasPlus[1], **population** denominators were obtained from U.S. Census county population estimates[6, 7], and county-level covariates captured social vulnerability and healthcare access/surveillance structure, including the **Social Vulnerability Index** (SVI; CDC/ATSDR) [8], **primary care physician density** from the Area Health Resources Files (AHRF) [9], **insurance coverage** derived from the SVI [8], **Health Professional Shortage Area (HPSA)**-derived healthcare capacity from the Health Resources and Services Administration (HRSA) [10], and **Federally Qualified Health Center (FQHC)** site density from HRSA [11]. **HIV diagnoses** and **gonorrhea counts** from CDC AtlasPlus were used for external validation[12, 13].The analytical workflow of the study is presented in Fig 1

**Fig 1.**
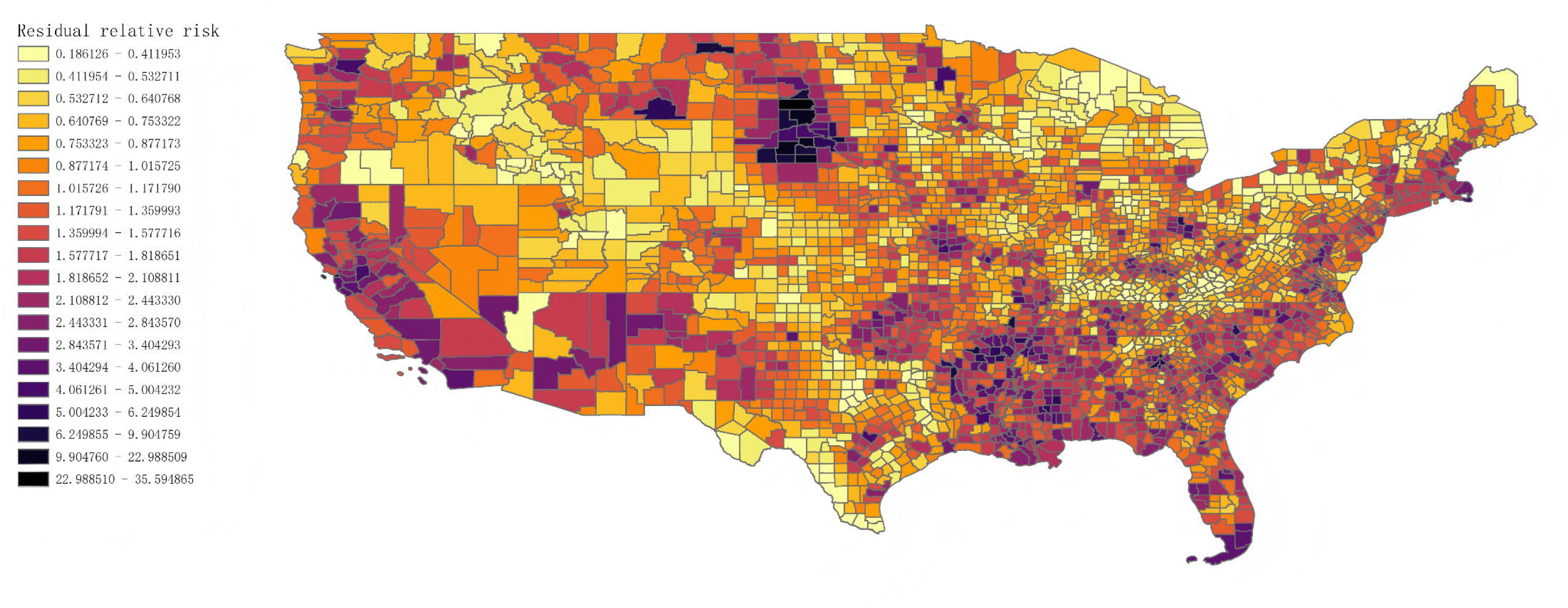
Analytical workflow of the study

### 2.1. Software

All statistical analyses were conducted in Python 3.13.13 using PyMC 5.26.1, ArviZ 0.22.0, pandas 2.3.3, NumPy 2.3.4, GeoPandas 1.1.2, libpysal 4.14.1, matplotlib 3.10.7, scikit-learn 1.8.0, statsmodels 0.14.6, and PyTensor 2.35.1. Final map figures were prepared in ArcGIS Pro 3.5.2.

### 2.2. Statistical model

We fit a Bayesian negative-binomial spatial model to annual county-level syphilis counts. Let *y_ij_* denote the observed number of primary and secondary syphilis cases in county *i* during year *j*, and let *µ_ij_* denote the corresponding expected count. The model was specified as

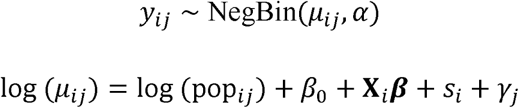

where pop_ij_ is the county-year population denominator included as an offset, *β*_0_ is the intercept, *x_i_* is the vector of standardized county-level covariates for county *i*, ***β*** is the corresponding vector of regression coefficients, s_i_ is the county-level spatial effect, *γ_j_* is the year-specific fixed effect for year *j*, and *α* is the negative-binomial overdispersion parameter.

The county-level spatial effect was further decomposed as

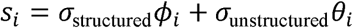

where *ø*_i_ is the structured spatial effect for county *i*, *θ*_*i*_ is the unstructured county-specific random effect, *σ*_structured_ is the scale parameter for the structured spatial component, and *a*_unstructured_ is the scale parameter for the unstructured spatial component. This formulation allowed the model to distinguish residual geographic clustering from non-spatial county-level heterogeneity while also accounting for year-to-year variation through *y*_*j*_[14].

### 2.3. Residual risk classification

#### 2.3.1. Posterior exceedance probability

From the fitted model, we defined the county-level residual relative risk as the exponentiated county spatial effect:

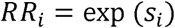

We then calculated the posterior exceedance probability

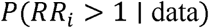

for each county[15]. Counties were grouped into three exceedance-probability tiers: Moderate (*≥0.80*), High (*≥0.90*), and Very high (*≥0.95*). In addition, we defined a binary stably high-risk county flag for counties with posterior exceedance probability *≥0.80* and posterior mean residual relative risk *≥1.50*.

#### 2.3.2. Residual risk magnitude

To summarize the magnitude of county-level residual risk, we also calculated the posterior mean residual relative risk,

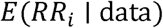

for each county. This quantity was used as the county-level risk magnitude measure.

### 2.4. Model diagnostics and sensitivity analyses

Model adequacy was assessed using several complementary diagnostic procedures. Convergence and sampler performance were evaluated using R-hat, effective sample size, divergent transitions, Bayesian fraction of missing information (BFMI), and tree depth. Approximate out-of-sample predictive performance was assessed using Pareto-smoothed importance sampling leave-one-out (PSIS-LOO) and Pareto-k diagnostics. Posterior predictive checks included observed-versus-predicted comparisons, zero-count calibration, and yearly aggregated totals. Residual spatial autocorrelation was evaluated using Moran’s I calculated on county-level mean Pearson residuals from the full fitted model. We also conducted a leave-one-covariate-out sensitivity analysis, in which each county-level covariate was omitted in turn while all other model specifications were held unchanged, to assess the robustness of the residual spatial risk estimates and high-risk county classifications.

### 2.5. External construct validation

To evaluate whether the county-level residual spatial signal identified by the main syphilis model corresponded to broader STI-related burden, we conducted an external construct validation analysis using HIV diagnoses and gonorrhea case counts. In the primary validation analysis, the posterior mean residual relative risk from the main 2010–2022 syphilis model was carried forward as a county-level continuous validation measure. We also performed a binary sensitivity analysis comparing counties with residual relative risk *≥ 1* versus *<1*.

Validation models were fit using generalized estimating equations with a negative-binomial family, exchangeable within-county correlation structure, and a log-population offset[16]. For county *i* in year *j*, the validation model can be written as

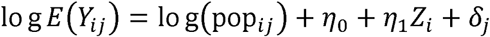

where *Y_ij_* denotes the HIV or gonorrhea count, *pop_ij_* is the county-year population denominator, *z_i_* is the county-level residual-risk measure derived from the syphilis model, *η*_0_ is the intercept, *η*_l_ is the association parameter of interest, and *δ_j_* represents year fixed effects. In the primary analysis, *z_i_* was the posterior mean residual relative risk; in the sensitivity analysis, *z_i_* was replaced by a binary indicator for counties with residual relative risk *≥ 1*. Associations were summarized as incidence rate ratios (IRRs) with 95% confidence intervals and p-values. The rationale for this validation step was that syphilis, HIV, and gonorrhea share overlapping transmission contexts and affected populations; therefore, counties with higher residual syphilis risk would be expected to show higher burdens of related STI outcomes.

## 3. Results

### 3.1. Descriptive summary

After harmonizing county identifiers across data sources, the final analytic panel included 3,109 counties in the contiguous United States and the District of Columbia, yielding 40,417 county-year observations from 2010 to 2022. Of these, 40,276 county-years had observed, non-suppressed primary and secondary syphilis counts and contributed to the main likelihood. Observed cases totaled 386,119, increasing from 13,627 in 2010 to 58,373 in 2022 (see Fig 2).

**Fig 2.**
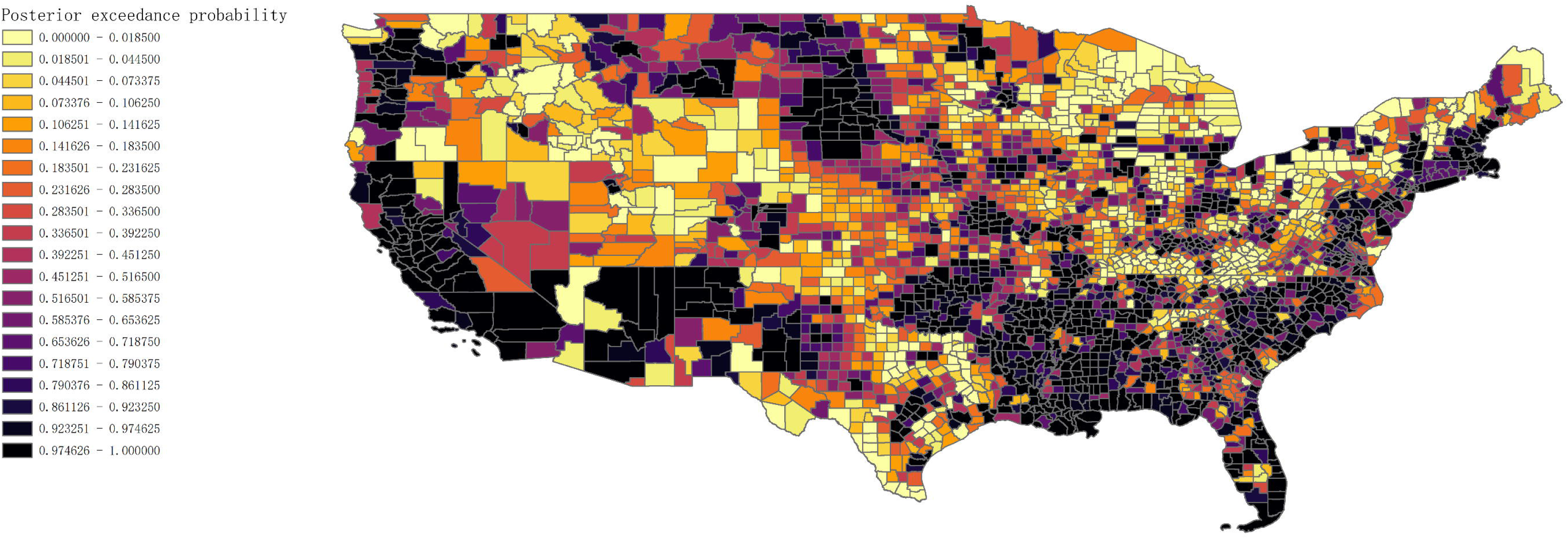
Annual observed primary and secondary syphilis cases in the contiguous United States and the District of Columbia, 2010–2022

Within the analytic syphilis panel, 24 county-years were reported as suppressed and 117 were unavailable, yielding 141 non-observed county-years in total. The corresponding gonorrhea panel contained 25 suppressed and 117 unavailable county-years, whereas HIV validation data contained 14,030 suppressed county-years.

The county-level adjustment covariates represented five categories of measurable surveillance and healthcare structure: social vulnerability, primary care physician density, insurance coverage, HPSA-derived healthcare capacity, and active FQHC site density. These variables were used as adjustment factors rather than causal exposures.

### 3.2. Model convergence and diagnostics

The main model showed good overall convergence. Sampling produced 0 divergent transitions, the maximum observed tree depth was 7, and no iterations reached the prespecified maximum tree depth. Across the primary model parameters, R-hat values were all at or below 1.016, and BFMI values ranged from 0.77 to 0.88, supporting adequate posterior exploration.

Posterior predictive performance was broadly reasonable for a sparse and overdispersed county-level count outcome. The observed proportion of zero counts was 53.2%, compared with a posterior predictive mean of 51.1%.

Remaining spatial autocorrelation in county-level mean Pearson residuals was small and not statistically significant (Moran’s I = 0.016; two-sided permutation p = 0.134), suggesting that the structured spatial component captured most county-level clustering after covariate adjustment.

PSIS-LOO diagnostics were interpreted cautiously because a small number of county-year observations were influential. Overall, 99.6% of observations had Pareto k values below 0.5, whereas 12 observations (0.03%) had Pareto k values greater than 1.0.

These influential observations were concentrated largely in rural and several tribal-area counties, mostly with relatively small populations, and therefore LOO-based summaries were treated as supplementary rather than primary evidence of model adequacy.

Yearly aggregate posterior predictive checks indicated a systematic temporal mismatch. Annual totals were underestimated in the early study years, were close to the observed totals around 2017, and became progressively overestimated from 2018 onward. Additional comparison of yearly fitted μ totals and yearly posterior predictive totals showed the same pattern, indicating that the late-period mismatch arose from the fitted mean structure rather than from posterior predictive variance alone. This issue is noted here and addressed further in the Supporting information S2 Text.

Leave-one-covariate-out sensitivity analyses(covariate) are summarized in Table 1. Omitting the social vulnerability index produced by far the largest deterioration in model fit (ΔLOOIC = +112.4) and the greatest departure from the main residual risk surface, indicating that it was the most influential adjustment variable in the model. Omitting primary care physician density had a smaller but still detectable effect on model fit and residual risk rankings, whereas omission of insurance coverage, HPSA-derived healthcare capacity, or FQHC site density produced only limited changes. Together, these results indicate that covariate adjustment was not redundant: the included covariates, especially social vulnerability and primary care physician density, materially corrected the observed spatial pattern and yielded a residual risk surface that more plausibly reflects underlying county-level risk.

**Table 1.**
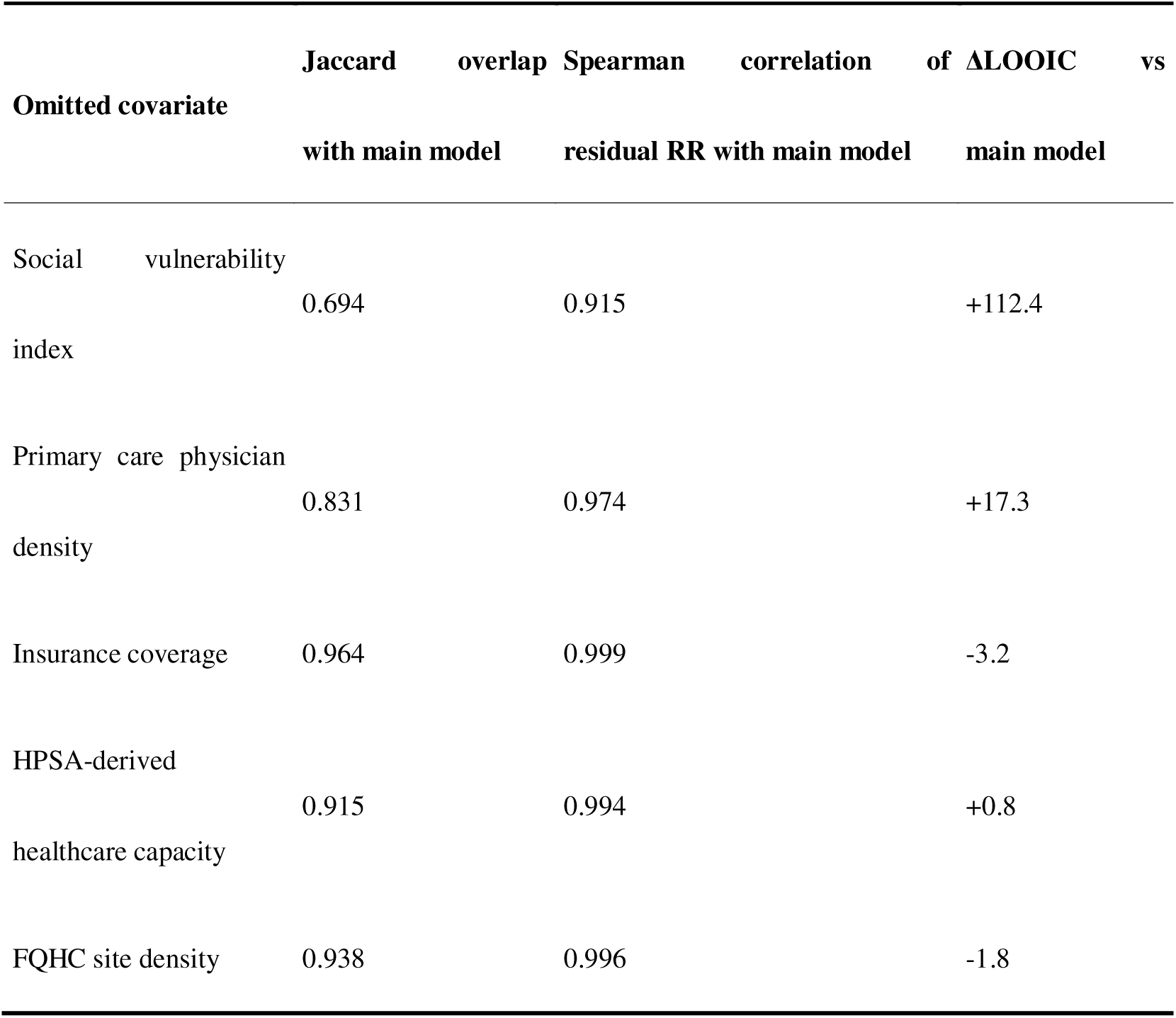
Leave-one-covariate-out sensitivity analysis of the residual spatial risk surface relative to the main model.

### 3.3. Fixed effects

The county-level covariates showed interpretable directional associations with reported syphilis burden. Higher social vulnerability and greater primary care physician density were positively associated with reported syphilis rates, whereas higher HPSA-derived healthcare capacity and greater FQHC site density were negatively associated with reported syphilis rates. The association with insurance coverage was weaker and may be more complex to interpret than the other covariate effects. Overall, the covariate patterns were directionally consistent with the study framework, and detailed posterior effect estimates are presented in Table 2.

**Table 2.** Covariate-specific posterior incidence rate ratios from the main model.

| Covariate | IRR | 95% CrI | P(IRR > 1) |
| --- | --- | --- | --- |
| Vulnerability index | 1.60 | 1.54–1.66 | 1.00 |
| Primary care physicians per 100,000 | 1.23 | 1.19–1.26 | 1.00 |
| Insurance rate | 1.04 | 1.00–1.09 | 0.98 |
| HPSA capacity | 0.91 | 0.88–0.94 | 0.00 |
| FQHC sites per 100,000 | 0.90 | 0.87–0.94 | 0.00 |

### 3.4. County-level residual spatial risk and posterior exceedance probability

#### 3.4.1. Residual spatial risk

Counties were first summarized according to posterior mean residual relative risk (RR). Across 3,109 counties, 1,527 had residual RR <1.0, 706 had residual RR between 1.0 and <1.5, 395 had residual RR between 1.5 and <2.0, 313 had residual RR between 2.0 and <3.0, and 168 had residual RR >=3.0 (see Supporting information S1 Table for more details). Higher residual spatial risk was concentrated primarily in the South, particularly in Mississippi, Georgia, Louisiana, Arkansas, Alabama, Florida, and North Carolina, with additional concentrations in California, Oklahoma, Missouri, Virginia, and parts of South Dakota (see Fig 3).

**Fig 3.**
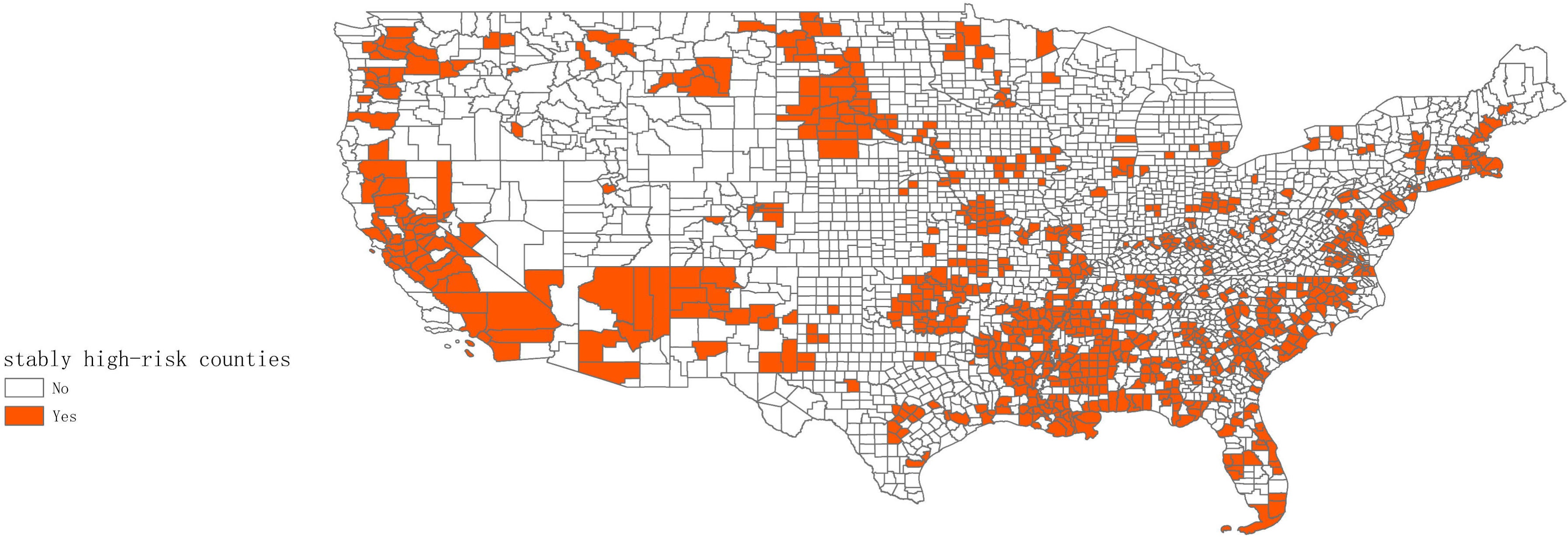
County-level residual relative risk the counties with the highest residual RR(top 10) are summarized in Table 3.

**Table 3.** Top 10 counties by residual relative risk.

| Rank | County | State | FIPS | Posterior mean residual RR |
| --- | --- | --- | --- | --- |
| 1 | Corson County | South Dakota | 46031 | 35.59 |
| 2 | Dewey County | South Dakota | 46041 | 22.99 |
| 3 | Todd County | South Dakota | 46121 | 15.37 |
| 4 | Jones County | South Dakota | 46075 | 13.56 |
| 5 | San Francisco County | California | 06075 | 13.12 |
| 6 | Jackson County | South Dakota | 46071 | 12.50 |
| 7 | Buffalo County | South Dakota | 46017 | 9.90 |
| 8 | Mellette County | South Dakota | 46095 | 9.14 |
| 9 | Oglala Lakota County | South Dakota | 46102 | 8.89 |
| 10 | Bennett County | South Dakota | 46007 | 8.61 |

#### 3.4.2. Posterior exceedance probability

Posterior exceedance probability was defined as P(RR_i > 1 | data), that is, the posterior probability that a county’s residual relative risk exceeded the null value of 1. Across 3,109 counties, 1,636 had exceedance probability <0.50, 424 had probabilities between 0.50 and <0.80, 236 had probabilities between 0.80 and <0.95, and 813 had probabilities >=0.95 (see Supporting information S1 Table for more details). Overall, 1,049 counties had exceedance probability >=0.80, indicating at least moderate posterior support for elevated residual risk, and 813 were in the very-high probability tier. High exceedance probabilities were concentrated primarily in the South, especially in Georgia, Mississippi, Louisiana, North Carolina, Arkansas, Alabama, Florida, Texas, Tennessee, and South Carolina, with additional clusters in Missouri, California, Oklahoma, Virginia, and parts of South Dakota (see Fig 4).

**Fig 4.**
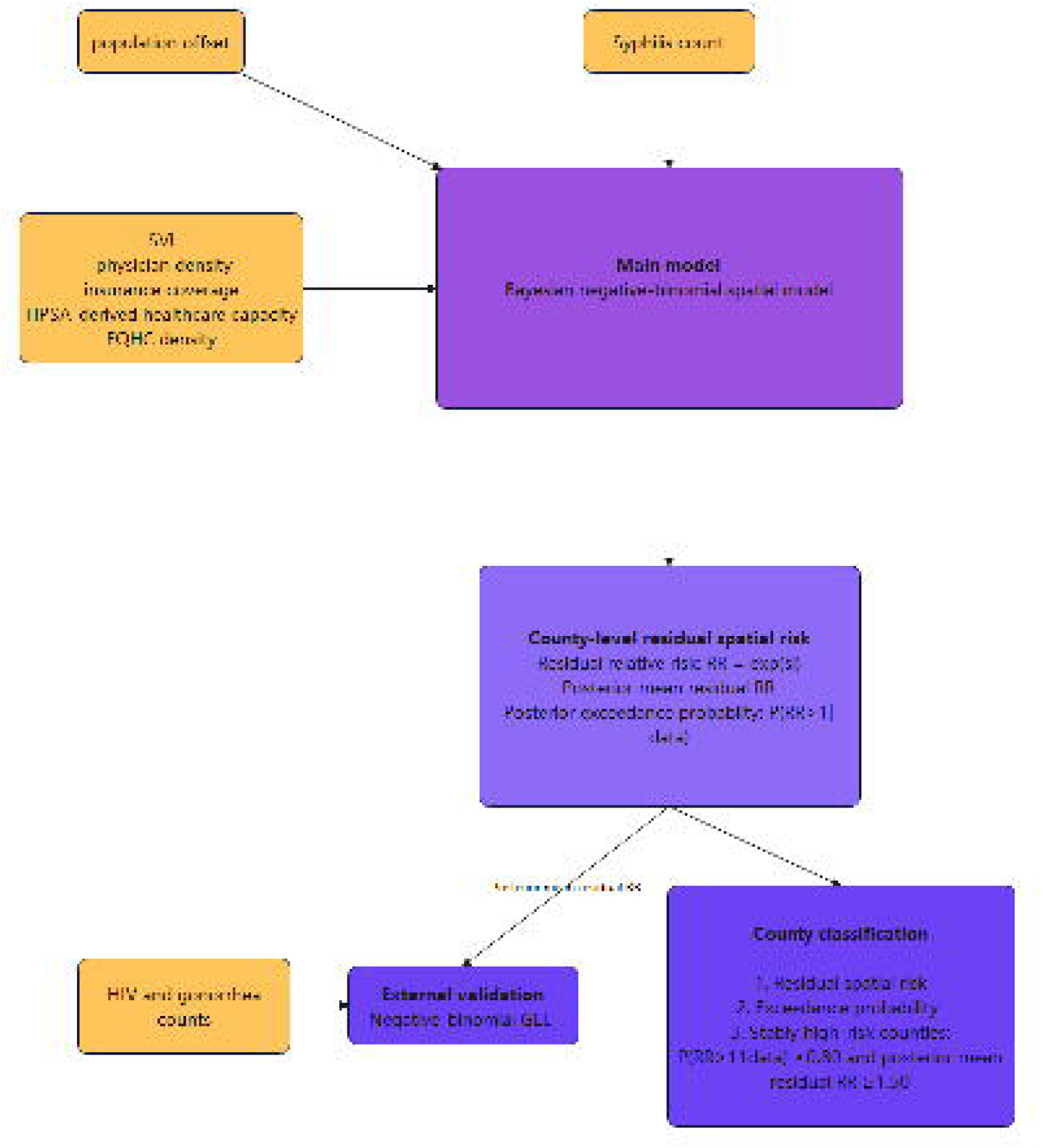
Posterior exceedance probability

#### 3.4.3. Stably high-risk counties

To identify stably high-risk counties, we defined a binary stably high-risk county flag as posterior exceedance probability *P(RR_*i*_ >1 1* data*) ≥0.80* together with posterior mean residual relative risk *≥1.50*. Under this definition, 850 counties were classified as stably high risk. These counties were concentrated primarily in the South, especially in Mississippi, Georgia, Louisiana, Arkansas, Alabama, Florida, North Carolina, South Carolina, Tennessee, and Texas, with additional clusters in Missouri, Oklahoma, Virginia, California, and South Dakota. The geographic distribution of these stably high-risk counties is shown in Fig 5.

**Fig 5.**
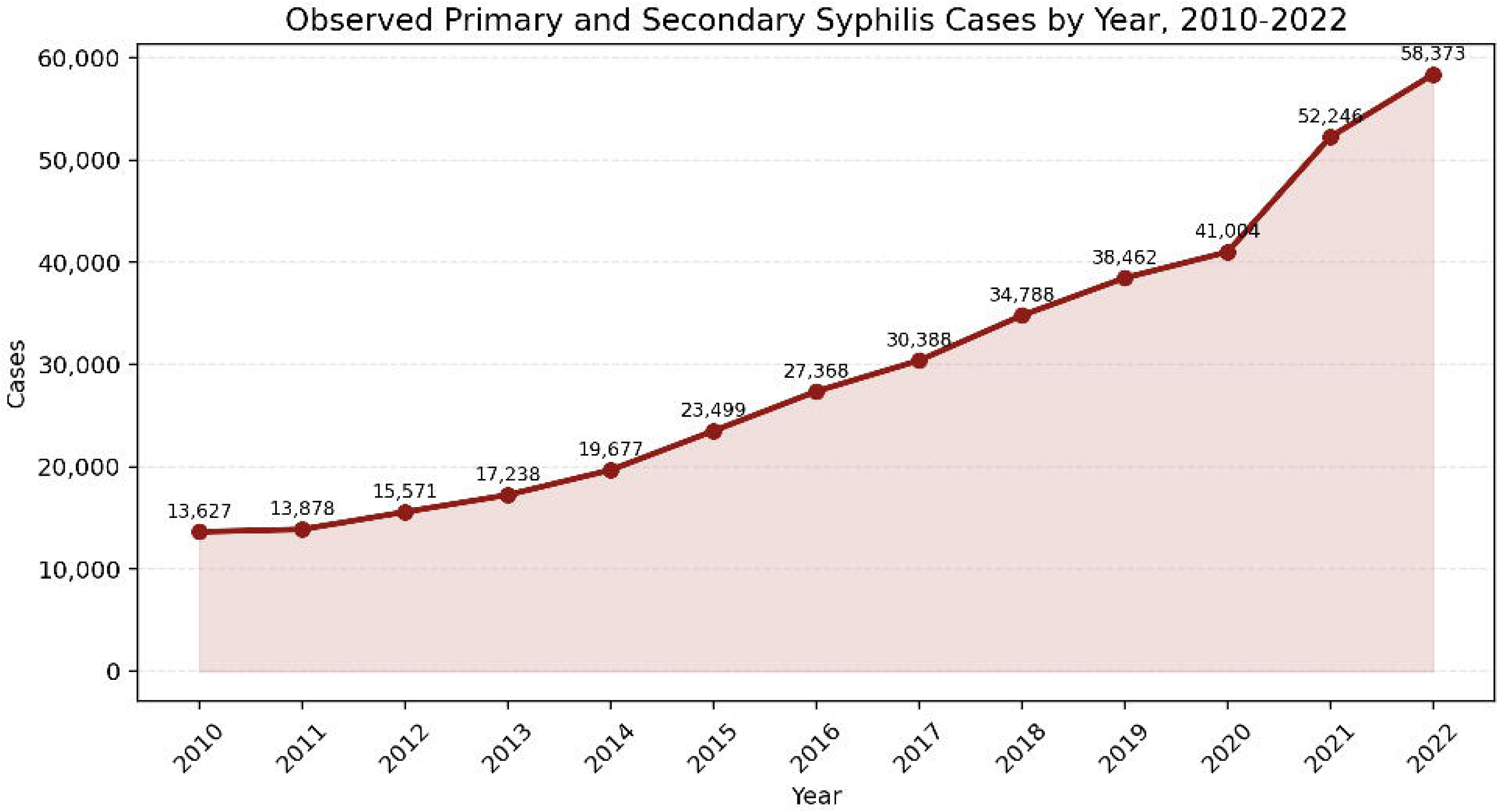
Geographic distribution of stably high-risk counties

### 3.5. External construct validation

Higher county-level residual syphilis risk from the main model was associated with higher HIV diagnosis rates (IRR = 1.63, 95% CI: 1.58–1.67, p = 3.55 × 10^-244^) and higher gonorrhea rates (IRR = 1.54, 95% CI: 1.51–1.58, p = 2.20 × 10^-274^). In sensitivity analyses, counties with residual relative risk greater than or equal to 1 also had higher HIV diagnosis rates (IRR = 2.87, 95% CI: 2.55–3.24, p = 8.39 × 10^-66^) and higher gonorrhea rates (IRR = 2.31, 95% CI: 2.15–2.48, p = 2.82 × 10^-123^) than counties with residual relative risk below 1. These findings support the external construct validity of the derived county-level residual risk surface and suggest that the identified spatial pattern was not unique to syphilis alone, but was meaningfully associated with the broader geographic distribution of STI risk in the contiguous United States. (see Table 4).

**Table 4.** External validation of main-model residual risk using HIV and gonorrhea outcomes.

| Analysis | Outcome | Residual-risk measure | IRR | 95% CI | p-value |
| --- | --- | --- | --- | --- | --- |
| External validation | HIV | Residual relative risk (per 1-unit increase) | 1.63 | 1.58–1.67 | 3.55e-244 |
| Sensitivity analysis | HIV | Higher residual-risk counties (RR ≥ 1) vs lower residual-risk counties (RR < 1) | 2.87 | 2.55–3.24 | 8.39e-66 |
| External validation | Gonorrhea | Residual relative risk (per 1-unit increase) | 1.54 | 1.51–1.58 | 2.20e-274 |
| Sensitivity analysis | Gonorrhea | Higher residual-risk counties (RR ≥ 1) vs lower residual-risk counties (RR < 1) | 2.31 | 2.15–2.48 | 2.82e-123 |

### 3.6. Spatial effect structure

The structured spatial component was substantially larger than the unstructured county-level component (sigma_structured = 1.46 vs sigma_unstructured = 0.03), indicating that residual variation in the model was dominated by geographically structured rather than spatially unstructured heterogeneity. This pattern is consistent with pronounced county-level clustering in syphilis risk and suggests that most residual spatial dependence was captured by the (ICAR) term.

Consistent with this interpretation, Moran’s I calculated on county-level Pearson residuals from the full fitted model was small and not statistically significant (Moran’s I = 0.016, two-sided permutation p = 0.134), indicating little remaining spatial autocorrelation after accounting for the modeled spatial structure (see supporting information S2 Text for more details).

## 4. Discussion

This study provides a national Bayesian spatial assessment of reported primary and secondary syphilis across all counties in the contiguous United States and the District of Columbia. After adjustment for measurable social vulnerability and healthcare- and surveillance-related structure, we identified 850 counties classified as stably high risk, indicating that substantial residual geographic heterogeneity remained even after accounting for observed county-level covariates. The strongest residual concentrations were observed primarily in the South, with additional clusters in selected western and midwestern counties. Importantly, the resulting county-level spatial signal was supported by external validation: counties identified as high priority or having higher residual risk also showed consistently higher HIV and gonorrhea burdens, indicating that the mapped residual risk pattern was not merely a model-derived numerical artifact, but was meaningfully associated with geographic syphilis risk and with the distribution of other sexually transmitted infections.

The covariate effects were broadly consistent with prior expectations, but they should be interpreted as adjustment terms rather than causal estimates. The strong positive association for the social vulnerability index was consistent with the well-established relationship between social deprivation and reported syphilis rates.The positive associations for primary care physician density and insurance coverage are more plausibly interpreted as reflecting surveillance and case-detection effects than as evidence of a direct causal increase in underlying risk. By contrast, the negative coefficients for FQHC site density and HPSA-derived healthcare capacity may indicate either lower reported rates in underserved areas because of under-detection, or a protective effect associated with greater healthcare access and service availability. Accordingly, the primary inferential target of this study is not the covariate coefficients themselves, but the residual spatial relative risk that remains after adjustment for these measurable county-level factors.

Compared with earlier local or regional spatial studies of syphilis, our analysis provides a national county-level view of geographic risk and therefore has greater relevance for national prioritization. At the national level, Chang et al. identified primary and secondary syphilis hot spots mainly in the South and Southeast, especially across the Gulf South, with additional hot spots in California and Maryland[17]. Although direct comparison should be made cautiously because that study used hot spot analysis of temporally smoothed 2008–2012 rates, whereas our study estimated adjusted residual spatial risk from a Bayesian model, our findings suggest a broader and more diffuse contemporary pattern. In addition to the persistent southern core, high-risk counties in our study extended more clearly along the Atlantic coastal corridor in a band-like pattern, and additional clusters were observed in parts of the north-central United States. Relative to this earlier literature, our study has three main advantages. First, county-level covariates were incorporated directly into risk estimation, rather than being examined only after hot spots had already been identified, thereby reducing potential distortion from measurable surveillance and healthcare-related structure. Second, rather than relying on a single cross-sectional hot spot map, we evaluated county-level hot spots (*RR > 1*) using posterior probabilities and further identified stably high-risk counties. Third, we summarized a set of stably high-risk counties, which provides a more robust picture of persistent geographic risk than conventional hotspot detection alone.

### 4.1. Limitations

This study has several limitations. First, its ecological design supports inference at the county level rather than at the level of individual risk. Second, the county-level covariates were treated as time-invariant and were derived from the most recent available cross-sections, so temporal changes in surveillance or healthcare capacity were not explicitly captured. Suppressed counts were excluded rather than imputed; in scenario checks, recoding all suppressed syphilis counts as 0, 1, or 3 changed the aggregate total by only 0–72 cases out of 386,119, suggesting minimal impact on overall case totals, although full refitted suppression sensitivity models were not performed. Fourth, posterior predictive yearly aggregates showed overestimation in later study years, likely reflecting the additive log-linear specification of spatial and temporal effects without a space-time interaction term. However, this issue primarily concerns temporal calibration rather than the main inferential target of the study, because our conclusions focus on county-level residual spatial risk classification rather than on formal prediction. Fifth, 12 observations (0.03%) had Pareto-k values greater than 1.0 and were concentrated largely in smaller-population counties, making substantial distortion of the overall county-level residual risk ranking unlikely. Finally, external validation used HIV and gonorrhea outcomes, which share important epidemiologic features with syphilis but do not perfectly overlap with syphilis transmission patterns.

### 4.2. Conclusions

Bayesian spatial residual risk mapping provides a principled, surveillance-adjusted approach to identifying priority counties for enhanced syphilis response. The counties classified as stably high risk were concentrated in the southeastern United States and along the Gulf Coast, with additional clusters in the north-central region and band-like distributions along both the Atlantic and Pacific coasts. By adjusting for measurable social and healthcare-related structure, the resulting residual spatial risk surface is less likely to reflect distortion from surveillance and access differences and more plausibly reflects underlying county-level geographic risk. External validation further showed that these classifications were not merely model-derived numerical outputs, but were meaningfully associated with the geographic distribution of STI risk. County-level residual risk estimates and posterior exceedance probabilities are summarized in Supporting information S1 Table.

## Data available statement

All raw data used in this study are publicly available from CDC AtlasPlus, the U.S. Census Bureau, CDC/ATSDR Social Vulnerability Index, and HRSA data resources. The processed analytic datasets, metadata, county adjacency files, statistical code, and data underlying the figures and tables are stored in a <u>private reviewer-access repository</u> during peer review. If the manuscript is accepted for publication, these materials will be made publicly available without access restrictions, and the final repository DOI or accession link will be provided at acceptance.

## Data Availability

The data used in this study were obtained from publicly available, aggregated, and de-identified sources. The original data sources are described in the manuscript. The processed data and related analysis materials are currently stored in a Google Drive folder and are available at the link provided below.

https://docs.google.com/document/d/1He8zWOHLe99ELCEdjG5bZ_WxO_LMjyRg/edit

## Acknowledgments

The authors acknowledge the Centers for Disease Control and Prevention AtlasPlus, the U.S. Census Bureau, the Health Resources and Services Administration, the CDC/ATSDR Social Vulnerability Index program, and the Area Health Resources Files for providing the publicly available data resources used in this study.

## Supporting information

**S1 Text. Detailed Bayesian model specification, prior distributions, and Markov chain Monte Carlo settings.**

**S2 Text. Model diagnostic results and supplementary interpretation.**

**S1 Table. County-level residual spatial risk estimates, posterior exceedance probabilities, and stably high-risk county classification.**

## Notes

### Competing Interest Statement

The authors have declared no competing interest.

### Author Declarations

This study used only publicly available, aggregated, and de-identified human health data that were openly available before the initiation of the study. County-level primary and secondary syphilis case counts, HIV diagnosis counts, and gonorrhea case counts were obtained from CDC AtlasPlus. County population denominators were obtained from the U.S. Census Bureau county population estimates. County-level covariates were obtained from publicly available CDC/ATSDR Social Vulnerability Index data, Area Health Resources Files, and HRSA data resources, including HPSA and FQHC-related data. No individual-level identifiable human participant data were used, and no data access request, screening, or ethics committee approval was required.

