## Supplementary Text S1 for "Surveillance-adjusted syphilis risk mapping across U.S. counties: a Bayesian spatial analysis with external validation against HIV and gonorrhea outcomes"

**Supporting information S1 Text.** **Detailed Bayesian model specification, prior distributions, and Markov chain Monte Carlo settings**

### 1.Likelihood

We modeled annual county-level primary and secondary syphilis case counts using a Bayesian negative-binomial spatial model. Let $y_{ij}$ denote the observed syphilis case count in county $i$ during year $j$, and let $\mu_{ij}$ denote the corresponding expected count. The outcome model was specified as

$$y_{ij}\sim\mathrm{NegBin}(\mu_{ij},\alpha)$$

where $y_{ij}$ is the observed county-year syphilis count, $\mu_{ij}$ is the expected county-year count, and $\alpha$ is the negative-binomial overdispersion parameter. We specified a negative-binomial likelihood rather than a Poisson likelihood to account for overdispersion in county-year syphilis counts(1).

### 2.Linear predictor

The logarithm of the expected count was modeled as

$$\log(\mu_{ij})=\log(\mathrm{pop}_{ij})+\beta_{0}+\mathbf{X}_{i}\boldsymbol{\beta}+s_{i}+\gamma_{j}$$

where $\mathrm{pop}_{ij}$ is the county-year population denominator included as an offset, $\beta_{0}$ is the intercept, $\mathbf{X}_{i}$ is the vector of county-level covariates for county $i$, $\boldsymbol{\beta}$ is the corresponding vector of regression coefficients, $s_{i}$ is the county-level spatial effect, and $\gamma_{j}$ is the year-specific fixed effect for year $j$. Thus, the model estimated expected syphilis counts as a function of population size, measured county-level covariates, spatially structured and unstructured residual variation, and year-to-year temporal differences.

#### 2.1.Population offset

The term $\log(\mathrm{pop}_{ij})$ is the natural logarithm of the county-year population denominator. Including this term as an offset means that the model is effectively estimating rates rather than raw counts, while preserving the count-data likelihood.

#### 2.2.Intercept

The intercept $\beta_{0}$ captures the baseline log-rate of syphilis after accounting for all other model components.

#### 2.3.County-level fixed effects

The covariate vector $\mathbf{X}_{i}$ included five county-level measures: social vulnerability, primary care physician density, insurance coverage, HPSA-derived healthcare capacity, and FQHC site density. These variables were treated as time-invariant in the main model. All covariates were standardized to zero mean and unit variance before model fitting so that regression coefficients were on a common scale and could be assigned comparable priors(2).

The coefficient vector $\boldsymbol{\beta}$ contains the fixed-effect regression coefficients corresponding to these standardized county-level covariates. Each coefficient quantifies the expected change in the log syphilis rate associated with a one-standard-deviation increase in the corresponding covariate, holding the other model components constant.

#### 2.4.County-level spatial effect

The county-level spatial effect was modeled as

$$s_{i}=\sigma_{\mathrm{structured}}\phi_{i}+\sigma_{\mathrm{unstructured}}\theta_{i}$$

where $s_{i}$ is the total spatial effect for county $i$, $\phi_{i}$ is the structured spatial component, $\theta_{i}$ is the unstructured county-specific random effect, $\sigma_{\mathrm{structured}}$ is the scale parameter for the structured spatial component, and $\sigma_{\mathrm{unstructured}}$ is the scale parameter for the unstructured component(3).

The structured term $\phi_{i}$ was implemented using an ICAR-type specification(4). In practice, the model first drew an unconstrained latent vector and then centered it so that the average structured spatial effect across counties was approximately zero. A pairwise penalty was then imposed across adjacent counties so that neighboring counties were encouraged to have similar values. This component therefore captures residual geographic clustering that is spatially smooth(4).

The unstructured term $\theta_{i}$ captures county-level heterogeneity that is not spatially patterned. Together, these two components allow the model to distinguish residual clustering from non-spatial county-to-county variability.

#### 2.5.Year effects

The term $\gamma_{j}$ represents the year-specific fixed effect for year $j$. Year effects were modeled as centered fixed effects rather than as a smooth random walk, allowing between-year variation to be estimated flexibly without imposing a specific smooth temporal trend. In implementation, a raw vector of year effects was assigned priors and then mean-centered so that the average year effect was approximately zero.

#### 2.6.Bayesian specification and prior distributions

From a Bayesian perspective, all unknown quantities in the model were treated as random variables. Prior distributions were assigned to the intercept, covariate coefficients, spatial scale parameters, overdispersion parameter, and raw year effects. Posterior inference was then obtained by combining these priors with the likelihood of the observed county-year syphilis counts.

To keep the model reasonably regularized while allowing the data to dominate, weakly informative priors were used. The intercept was assigned a wider prior because baseline syphilis rates may vary substantially on the log scale, whereas standardized covariate coefficients were assigned unit-scale normal priors. Conservative half-normal priors were used for the spatial scale parameters to discourage unrealistically large residual spatial variance while still allowing meaningful clustering to emerge. The overdispersion parameter was also assigned a half-normal prior.

Table 1 Prior distributions used in the main Bayesian spatial model

| **Parameter** | **Prior** | **Dimension** | **Description** |
| --- | --- | --- | --- |
| $\beta_{0}$ | $\mathcal{N}(0,2)$ | 1 | Model intercept |
| $\beta_{k}$ | $\mathcal{N}(0,1)$ | 5 | Standardized county-level covariate coefficients |
| $\sigma_{\mathrm{structured}}$ | HalfNormal$\left( 0.5 \right)$ | 1 | Scale of structured spatial component |
| $\sigma_{\mathrm{unstructured}}$ | HalfNormal$\left( 0.5 \right)$ | 1 | Scale of unstructured spatial component |
| $\theta_{i}$ | $\mathcal{N}(0,1)$ | 3,109 counties | Unstructured county-level random effects |
| $\phi_{i}^{\mathrm{raw}}$ | $\mathcal{N}(0,1)$ | 3,109 counties | Raw latent values used to construct the centered structured spatial effect |
| $\alpha$ | HalfNormal$\left( 2 \right)$ | 1 | Negative-binomial overdispersion parameter |
| $\gamma_{j}^{\mathrm{raw}}$ | $\mathcal{N}(0,0.5)$ | 13 years | Raw year effects before centering |

In the absence of strong substantive prior information for this application, the main model relied primarily on weakly informative priors so that the data could play the dominant role in posterior inference(2). Specifically, weakly informative normal priors were assigned to the model intercept and standardized regression coefficients $\beta_{0}$and $\beta_{k}$(2). Similarly, weakly informative regularizing priors were assigned to the parameters that govern the magnitude of the spatial, temporal, and dispersion components, including the structured and unstructured spatial scale parameters $\sigma_{\text{structured}}$and $\sigma_{\text{unstructured}}$, the raw year effects $\gamma_{j}^{\text{raw}}$, and the negative-binomial overdispersion parameter $\alpha$(2). Standard normal priors were used for the latent components entering the spatial random effects parameterization $\varphi_{i}^{\text{raw}}$and $\theta_{i}$, while the intrinsic conditional autoregressive (ICAR) potential imposed the structured spatial smoothing constraint(5). Together, these priors were intended to stabilize estimation and discourage implausibly large spatial, temporal, or dispersion components without imposing strong substantive prior assumptions.

Because regularizing priors on spatial, temporal, and scale parameters can in principle influence posterior smoothing, county-level residual risk estimation, and the identification of stably high-risk counties, a prior sensitivity analysis was conducted.

The model was refit under both wider weakly informative priors and moderately stronger regularizing priors while all non-prior aspects of the analysis were held unchanged.

The resulting county-level residual risk surface, posterior probabilities that county-level residual risk exceeded 1, and stably high-risk county classifications were highly stable across prior specifications, indicating that the main conclusions were not materially driven by the prior settings (see S2 Text).

#### 2.7.Derived county-level quantities

the county-level residual spatial risk was defined on the relative-risk scale as the exponentiated county spatial effect,

$$RR_{i}=\exp(s_{i})=\exp(\sigma_{\mathrm{structured}}\phi_{i}+\sigma_{\mathrm{unstructured}}\theta_{i})$$

where $RR_{i}$ represents the residual county-level relative risk after accounting for the population offset, measured covariates, and year effects. For each posterior draw, this quantity was calculated by exponentiating the fitted county spatial effect. The reported county-level summary residual_rr_mean corresponds to the posterior mean of $RR_{i}$ across retained posterior draws, whereas log_rr_spatial_mean corresponds to the posterior mean of the county spatial effect on the log scale.

$$P(RR_{i}>1\mid\mathrm{data})$$

which quantifies the posterior probability that the residual relative risk for county $i$ exceeds the null value of 1(6).

A binary high-priority county flag was additionally defined for counties with posterior exceedance probability $\geq0.80$ and posterior mean residual relative risk $\geq1.50$. This definition was used to identify counties with both substantial residual risk magnitude and reasonably stable posterior support.

#### 2.8.Spatial adjacency structure

County adjacency was defined using Queen contiguity based on the 2022 TIGER/Line county shapefile. Two counties were considered adjacent if they shared either a boundary segment or a vertex. This adjacency structure was used to construct the neighborhood graph for the structured spatial component.

#### 2.9.MCMC sampling configuration

Posterior sampling was performed in PyMC using the No-U-Turn Sampler (NUTS)(7, 8). Four chains were run, each with 2,000 tuning iterations and 2,000 retained posterior draws. The target acceptance probability was set to 0.99, the maximum tree depth was set to 14, and the random seed was 42. The posterior sampling configuration used for the main Bayesian spatial model is summarized Table 2.

Table 2 Posterior sampling configuration

| **Parameter** | **Value** |
| --- | --- |
| Number of chains | 4 |
| Tuning iterations per chain | 2,000 |
| Retained posterior draws per chain | 2,000 |
| Total retained posterior draws | 8,000 |
| Target acceptance probability | 0.99 |
| Maximum tree depth | 14 |
| Random seed | 42 |

### 3.External validation model

External validation was conducted using HIV diagnoses and gonorrhea case counts as related STI outcomes. In the primary external validation analysis, the posterior mean county-level residual relative risk from the main 2010–2022 syphilis model was used as a continuous county-level validation measure. A binary sensitivity analysis was also performed by comparing counties with residual relative risk $\geq1$ versus counties with residual relative risk $\left< 1 \right.$.

For county $i$ in year $j$, the external validation model was specified as

$$\mathrm{lo}g E\left( Y_{ij} \right)=\mathrm{lo}g \left( \mathrm{pop}_{ij} \right)+\eta_{0}+\eta_{1}Z_{i}+\delta_{j}$$

where $Y_{ij}$ denotes the HIV or gonorrhea count, $\mathrm{pop}_{ij}$ is the county-year population denominator, $\eta_{0}$ is the intercept, $Z_{i}$ is the county-level residual-risk measure derived from the syphilis model, $\eta_{1}$ is the association parameter of interest, and $\delta_{j}$ represents year fixed effects. In the primary analysis, $Z_{i}$ was the continuous posterior mean residual relative risk; in the sensitivity analysis, $Z_{i}$ was a binary indicator for counties with residual relative risk $\geq1$. These models were fit using generalized estimating equations with a negative-binomial family, exchangeable within-county correlation structure, and a log-population offset(9). Associations were summarized as incidence rate ratios with 95% confidence intervals and p-values.

### 4.Model checking

Several classes of diagnostics were used to assess model adequacy. Convergence and sampler behavior were evaluated using R-hat, effective sample size, divergent transitions, BFMI, and tree depth. Approximate out-of-sample predictive performance was assessed using PSIS-LOO and Pareto-k diagnostics. Posterior predictive checks included observed-versus-predicted comparisons, zero-count calibration, and yearly aggregated totals. Residual spatial autocorrelation was evaluated using Moran’s I calculated on county-level mean Pearson residuals from the full fitted model. In addition, leave-one-covariate-out sensitivity analyses were performed to assess the contribution of each county-level covariate to adjustment of the residual spatial risk surface. Prior sensitivity analyses were also conducted by refitting the model under alternative prior scenarios while holding all non-prior aspects of the analysis fixed. Detailed interpretation of these diagnostic results is provided in Supporting information S2 Text.
