## Supplementary Text S2 for "Surveillance-adjusted syphilis risk mapping across U.S. counties: a Bayesian spatial analysis with external validation against HIV and gonorrhea outcomes"

**S2 Text. Model diagnostic results and supplementary interpretation**

### 1.Divergent Transitions

**What it tests:** Divergent transitions are a diagnostic specific to Hamiltonian Monte Carlo (HMC) samplers(1). During the numerical simulation of Hamiltonian dynamics, the integrator may encounter regions of the posterior geometry where the curvature changes sharply (e.g., funnel-shaped hierarchical posteriors). When this happens, the numerical trajectory diverges from the true Hamiltonian path, producing biased samples that may systematically avoid portions of the posterior(1). A divergent transition is therefore a direct indicator that the sampler may have failed to fully explore the target distribution (Betancourt, 2017; Stan Development Team, 2024).

**Classification thresholds:** Any divergent transition is potentially problematic. In practice, zero divergences is the target. A small number (e.g., < 0.1% of transitions) may be tolerable if other diagnostics are satisfactory, but even one divergence warrants investigation because it signals a region of the posterior that was not adequately explored(1).

**Current result:** 0 divergent transitions across all 4 chains. This confirms that the NUTS sampler successfully navigated the posterior geometry without encountering pathological curvature, and the posterior samples are not systematically biased by integration errors.

### 2.Maximum Tree Depth

**What it tests:** The No-U-Turn Sampler (NUTS) adaptively selects the length of the Hamiltonian trajectory by building a binary tree of leapfrog steps(2). A maximum tree depth cap is imposed to prevent excessively long trajectories. When iterations frequently reach this cap, NUTS is being prematurely terminated before the trajectory has fully explored the local posterior geometry, resulting in inefficient (though not invalid) sampling(2). Unlike divergences, hitting maximum tree depth is an efficiency concern rather than a validity concern (Hoffman and Gelman, 2014).

**Classification thresholds:** The prespecified maximum tree depth in our model was 14 (corresponding to 2^14^ - 1 = 16,384 leapfrog steps per iteration). If a substantial fraction of iterations hit this cap, the effective sample size per unit of computation time is reduced. The default in most PyMC implementations is 10(3); we increased it to 14 to provide ample headroom for the high-dimensional spatial random effects.

**Current result:** Maximum observed tree depth was 7 across all chains, and 0 iterations reached the prespecified maximum of 14. This indicates that the sampler efficiently explored the posterior without requiring excessively long trajectories. The low observed tree depth suggests that the sampler did not require long trajectories to explore the posterior under the fitted parameterization.

### 3.Split R-hat (Potential Scale Reduction Factor)

**What it tests:** The split R-hat statistic assesses whether multiple MCMC chains have converged to the same stationary distribution. Each chain is split in half, and the between-segment variance is compared to the within-segment variance. If chains have converged, these variances should be approximately equal, yielding R-hat ≈ 1.0. Values substantially greater than 1 indicate that the chains have not yet mixed — that is, different chains (or different portions of the same chain) are exploring different regions of the posterior(4).

**Classification thresholds:** The original recommendation by Gelman and Rubin (1992) was R-hat < 1.1(5). Vehtari et al. (2021) proposed a rank-normalized version of R-hat and tightened the recommendation to R-hat < 1.01(6). Our model uses 4 chains, and we adopt the stricter 1.01 threshold as the primary benchmark, with R-hat < 1.05 as a secondary "acceptable" criterion.

Current result: All primary model parameters had R-hat ≤ 1.016. The maximum observed R-hat (1.016 for sigma_unstructured) slightly exceeds the 1.01 threshold. However, sigma_unstructured had a posterior mean of 0.029, much smaller than the structured spatial standard deviation, suggesting that the unstructured county-level component contributed little additional variation beyond the ICAR spatial effect. This parameter was therefore close to the lower boundary of its parameter space, which may explain the slightly elevated R-hat (see Figure S1 for more details).

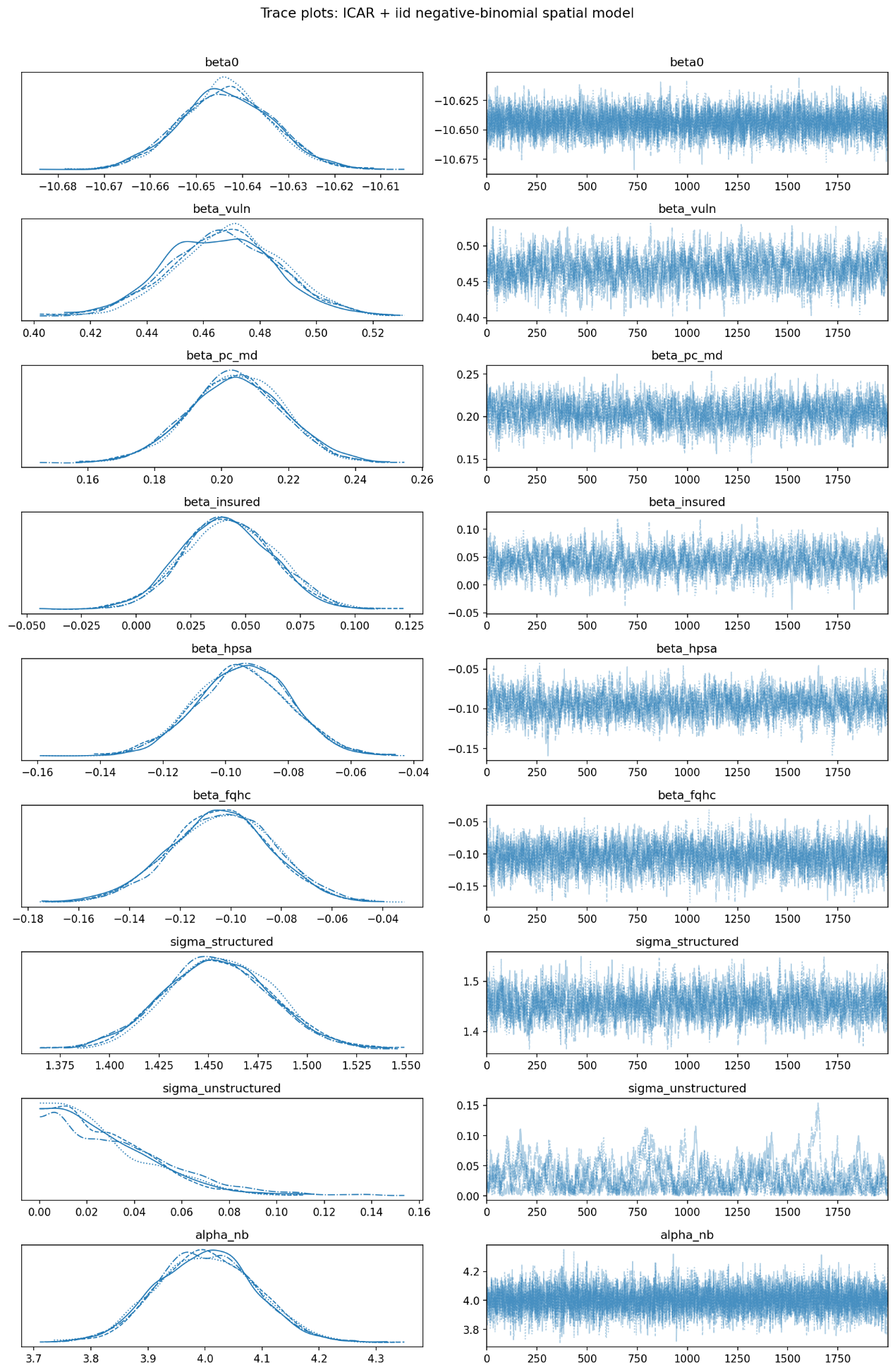

Figure S1 MCMC trace plots

Note: Trace plots of selected fixed effects, variance components, and the negative-binomial dispersion parameter across four MCMC chains. Most parameters showed stable mixing; sigma_unstructured was close to zero and showed greater relative fluctuation, consistent with its slightly elevated R-hat.

### 4.Effective Sample Size (ESS)

**What it tests:** Because MCMC draws are autocorrelated, the nominal number of posterior draws overstates the amount of independent information available for inference(7). Effective sample size (ESS) estimates how many independent draws would provide the same amount of information as the correlated MCMC output. Low ESS implies greater Monte Carlo uncertainty in posterior summaries(6). Bulk-ESS assesses sampling efficiency for central location summaries such as posterior means and medians, whereas tail-ESS assesses sampling efficiency for extreme quantiles and interval estimates(6, 7).

**Classification thresholds:** Stan currently recommends requiring bulk-ESS greater than 100 times the number of chains for final inference. With 4 chains, this corresponds to a recommended threshold of at least 400(7). Values below this threshold do not automatically invalidate the whole model, but they indicate that Monte Carlo error may still be non-negligible for the affected parameter and that parameter-specific inference should be interpreted cautiously.

**Current result:** In the main model, most parameters had bulk-ESS and tail-ESS comfortably above 400, indicating adequate sampling efficiency for the principal inferential targets. For example, the regression coefficients, sigma_structured, and alpha_nb all had bulk-ESS values above 900, with much larger tail-ESS values. The main exception was sigma_unstructured, which had the lowest bulk-ESS (293.0) and tail-ESS (376.5), both below the recommended threshold. This indicates that Monte Carlo precision for the unstructured variance component is weaker than for the rest of the model. Consistent with this, sigma_unstructured also had the largest R-hat in the table (1.0154). At the same time, its posterior mean was very small (0.0289) and its interval extended down to 0, suggesting that the unstructured county-level variance was weakly identified and close to the boundary of zero. Thus, ESS diagnostics support reliable inference for the main regression and structured spatial parameters, while inference for sigma_unstructured itself should be treated more cautiously.

Although the ESS and R-hat diagnostics for sigma_unstructured appear less favorable, this does not materially weaken the main model results. This parameter represents the unstructured county-level heterogeneity, which was estimated to be very small compared with the structured ICAR component (sigma_structured = 1.46 vs. sigma_unstructured = 0.029). In other words, almost all residual county-level spatial variation was captured by the ICAR component, while the unstructured component contributed little to the fitted risk surface. Therefore, the weak sampling diagnostics for this near-zero parameter are unlikely to affect the main fixed-effect estimates, residual risk maps, or posterior exceedance probabilities(see Table S1 for more details).

Table S1 Posterior summaries and MCMC diagnostics for main model parameters

| **Parameter** | **Description** | **Mean** | **SD** | **95% CrI** | **MCSE mean** | **MCSE SD** | **Bulk ESS** | **Tail ESS** | **R-hat** |
| --- | --- | --- | --- | --- | --- | --- | --- | --- | --- |
| β₀ (beta0) | Intercept | -10.644 | 0.010 | -10.663, -10.625 | 0.0001 | 0.0001 | 4,767 | 5,708 | 1.001 |
| β_vuln (beta_vuln) | Social vulnerability index | 0.468 | 0.019 | 0.430, 0.504 | 0.0006 | 0.0003 | 1,002 | 2,067 | 1.007 |
| β_pc_md (beta_pc_md) | Primary care physician density | 0.204 | 0.014 | 0.178, 0.231 | 0.0004 | 0.0002 | 1,347 | 2,695 | 1.003 |
| β_insured  (beta_insured) | Insurance coverage | 0.042 | 0.021 | 0.006, 0.083 | 0.0007 | 0.0003 | 946 | 1,830 | 1.003 |
| β_hpsa  (beta_hpsa) | HPSA-derived healthcare capacity | -0.095 | 0.015 | -0.124, -0.068 | 0.0004 | 0.0002 | 1,342 | 2,339 | 1.002 |
| β_fqhc  (beta_fqhc) | Active FQHC site density | -0.104 | 0.020 | -0.141, -0.065 | 0.0004 | 0.0002 | 2,150 | 3,590 | 1.002 |
| σ_structured  (sigma_structured) | Structured ICAR spatial SD | 1.455 | 0.027 | 1.405, 1.506 | 0.0008 | 0.0004 | 1,268 | 2,487 | 1.005 |
| σ_unstructured  (sigma_unstructured) | Unstructured county-level SD | 0.029 | 0.023 | 0.000, 0.068 | 0.0014 | 0.0013 | 293 | 376 | 1.015 |
| α_NB  (alpha_nb) | Negative-binomial dispersion | 4.002 | 0.085 | 3.851, 4.166 | 0.0009 | 0.0009 | 9,818 | 6,681 | 1.000 |

### 5.Bayesian Fraction of Missing Information (BFMI)

**What it tests:** BFMI is a diagnostic specific to HMC that evaluates how efficiently the momentum resampling step explores the energy levels of the posterior(8). It is computed as the ratio of the variance of successive energy transitions to the marginal variance of the energy(9). Low BFMI indicates poor exploration of the energy distribution and may suggest inefficient HMC sampling.

**Classification thresholds:** The current convention is that BFMI values below 0.3 indicate problematic sampling. This threshold is described as "provisional" in the ArviZ and Stan documentation, but it is widely used in practice (Betancourt, 2016; Stan Development Team, 2024). Values well above 0.3 (approaching 1.0) indicate excellent energy-level exploration.

Current software guidance commonly treats BFMI values below 0.3 as concerning. ArviZ states that values smaller than 0.3 indicate poor sampling, while noting that this threshold is provisional and may change(10); Stan likewise uses a nominal threshold of 0.3 and warns that HMC may have trouble exploring the target distribution when BFMI falls below it(7). Conceptually, higher BFMI values indicate more effective exploration of the marginal energy distribution, with BFMI approaching 1 representing the theoretical ideal(9).

**Current result:** Current result: BFMI values across the 4 chains were 0.84, 0.88, 0.77, and 0.81 (range: 0.77-0.88), all well above the nominal 0.3 threshold. These values indicate adequate exploration of the energy distribution by the HMC sampler and are consistent with the absence of divergent transitions and the low observed tree depths.

### 6.Posterior Predictive Check: Zero-count Proportion

**What it tests:**

A useful posterior predictive check for sparse county-level count data is whether the model reproduces the observed proportion of zero counts(11). In the Bayesian workflow, posterior predictive checks compare important features of the observed data with the same features computed from replicated data generated under the fitted model; discrepancies can indicate aspects of the data structure that the model fails to capture and may motivate model extension or refinement(12).

**Classification thresholds:** The comparison is primarily qualitative: the posterior predictive distribution of the zero proportion should be broadly consistent with the observed zero proportion. In practice, large discrepancies may warrant further investigation, but no universal numeric cutoff is established for this check.

**Current result:** Observed zero proportion = 53.2%; posterior predictive mean zero proportion = 51.1%. The 2.1 percentage point difference is small, indicating that the negative binomial likelihood with county-level random effects adequately captures the zero-generating process without requiring a separate zero-inflation component.

### 7.Residual Spatial Autocorrelation (Moran's I)

**What it tests:**

Moran’s I is a widely used statistic for assessing overall spatial autocorrelation across a study region, that is, whether nearby geographic units tend to have more similar values than would be expected under spatial randomness(13).

When applied to model residuals, it serves as a post-fit diagnostic for remaining spatial structure. Residual spatial autocorrelation is an important post-fit diagnostic for models fitted to spatial data. After fitting a spatial model, residual spatial autocorrelation should be substantially reduced; if it remains statistically detectable, this suggests that some spatial dependence may still be unaccounted for by the model, or that relevant spatially structured covariates or correlation structures have been omitted(14).

**Classification thresholds:**

Under the null hypothesis of no spatial autocorrelation, the expected value of Moran’s I is E[I] = -1/(n-1), which approaches 0 as n becomes large(15). Statistical significance was assessed using a Monte Carlo permutation test with 999 permutations, with p < 0.05 taken to indicate statistically significant spatial autocorrelation(15). A non-significant p-value indicates that the null hypothesis cannot be rejected and therefore provides no evidence of residual spatial autocorrelation.

**Current result:** Moran's I = 0.016, two-sided permutation p = 0.134. The residual spatial autocorrelation is small and not statistically significant, confirming that the ICAR structured spatial effect has adequately captured county-level geographic clustering. This is consistent with the very high spatial variance contribution (σ_structured = 1.46 >> σ_unstructured = 0.03).

**Suggested figure:** Figure S3 — Histogram of Moran's I values from the permutation distribution, with observed value marked.

### 8.PSIS-LOO and Pareto k Diagnostics

**What it tests:** What it tests: PSIS-LOO is a computational method for estimating out-of-sample predictive performance without refitting the model once per observation. In leave-one-out cross-validation, each observation is evaluated using the posterior predictive density obtained from a model fit to all of the other observations. PSIS approximates these leave-one-out quantities from the full posterior draws by stabilizing the corresponding importance weights(16). The Pareto k diagnostic is used to assess the reliability of the PSIS estimate(16). It is not itself a measure of predictive fit. Rather, it is a diagnostic of the stability and finite-sample behavior of the importance-sampling procedure: larger k values indicate that the corresponding PSIS estimate is less reliable and may be unstable or biased(16).

**Classification thresholds(17):**

Table S2 presents the Pareto k̂ classification thresholds used to assess the reliability of PSIS-LOO estimates.

Table S2 Pareto k̂ diagnostic classification thresholds

| **Pareto (k) range** | **Interpretation** |
| --- | --- |
| $\left( k<\min\left( 1-\frac{1}{\log_{10} \left( S \right)}, 0.7 \right) \right)$ | The PSIS estimate and the corresponding Monte Carlo standard error estimate are reliable. |
| $\left( 1-\frac{1}{\log_{10} \left( S \right)}\leq k<0.7 \right)$ | The PSIS estimate and the corresponding Monte Carlo standard error estimate are not reliable, but increasing the effective sample size (S) above 2200 may help. |
| $\left( 0.7\leq k<1 \right)$ | The PSIS estimate and the corresponding Monte Carlo standard error have large bias and are not reliable. |
| $\left( k\geq1 \right)$ | The target distribution is estimated to have a non-finite mean. The PSIS estimate and the corresponding Monte Carlo standard error are not well defined. |

Note: In the present analysis, the posterior sample size was S = 8,000 (4 chains × 2,000 post-warmup draws), yielding a sample-size-dependent threshold of $1-\frac{1}{\log_{10} \left( S \right)}, 0.7$ = 0.744. Because this exceeds 0.7, the effective threshold reduces to min(0.744, 0.7) = 0.7, and the second tier in Table S2 does not apply.

**Current result:** Of 40,276 total observations, 40,112 (99.59%) had Pareto k < 0.5, indicating favorable PSIS-LOO reliability for the large majority of observations. Only 12 observations (0.03%) had k > 1.0. These high-k observations were concentrated largely in rural and several tribal-area counties, mostly with relatively small populations (e.g., Oglala Lakota County, SD; Dewey County, SD; Todd County, SD). In such settings, sparse counts and small population denominators can produce observations that are difficult to approximate well under leave-one-out importance sampling. This pattern is not unusual in county-level disease mapping and does not by itself imply broad model failure（see Figure S2）.

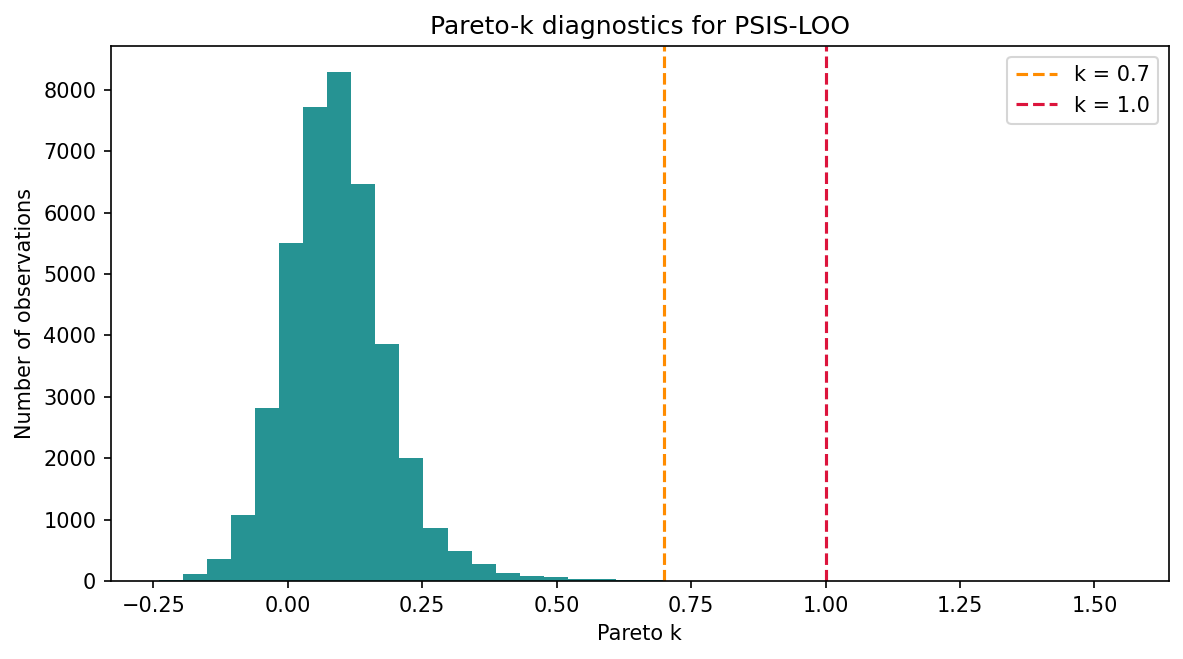

Figure S2 Distribution of Pareto k diagnostics from PSIS-LOO for the main 2010–2022 model

### 9.Yearly Aggregate Posterior Predictive Check

**What it tests:** Posterior predictive checks (PPCs) assess whether data replicated under the fitted model resemble the observed data in relevant features(12). Observation-level posterior predictive checks examined whether the fitted model reproduced the overall distribution of county-year case counts(see Figure S3);whereas yearly aggregate posterior predictive checks examined whether annual totals summed across counties reproduced the observed year-to-year temporal pattern.

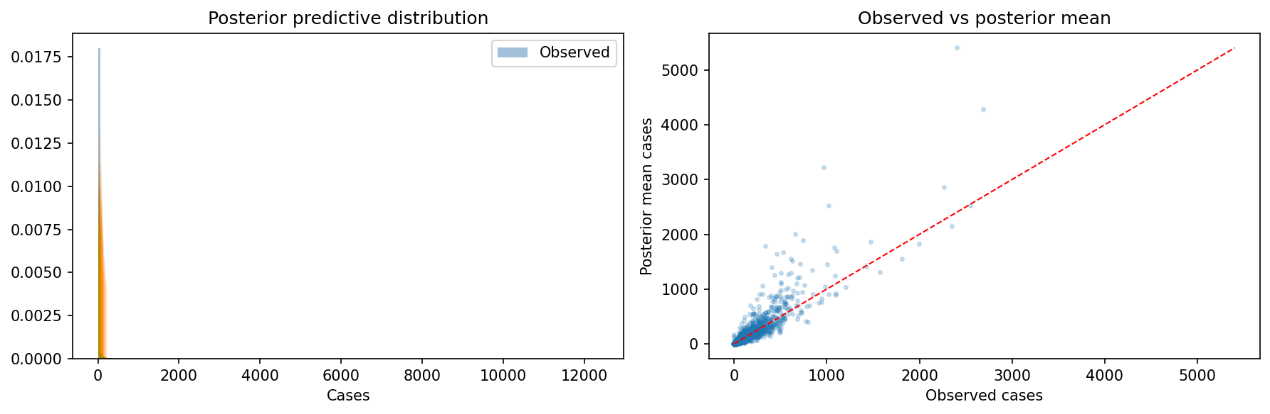

Figure S3 Observation-level posterior predictive checks for the main 2010–2022 model, showing the observed county-year syphilis count distribution alongside posterior predictive replicated distributions (left) and observed versus posterior mean county-year counts (right).

**Classification thresholds:** There are no fixed thresholds. The assessment is whether the predicted yearly totals track the observed totals within the posterior predictive uncertainty interval. Systematic bias (consistently over- or under-predicting across multiple years) is more concerning than random scatter.

**Current result:** Annual posterior predictive totals underestimated observed counts in the early study years (2010–2013), matched observed counts around 2015–2017, and progressively overestimated them in later years (2020–2022). In 2022, the posterior predictive annual total was 1.62 times the observed total (94,746.8 vs. 58,373 cases). An additional diagnostic comparing yearly totals from the fitted mean (μ) versus from posterior predictive draws (y_rep) showed identical patterns, confirming that this mismatch arises from the fitted mean structure, not from negative binomial sampling variance. Because yearly mu_ratio and ppc_ratio were nearly identical, the annual mismatch was already present in the fitted mean structure before negative-binomial sampling was applied（see Table S3 and for more details） .

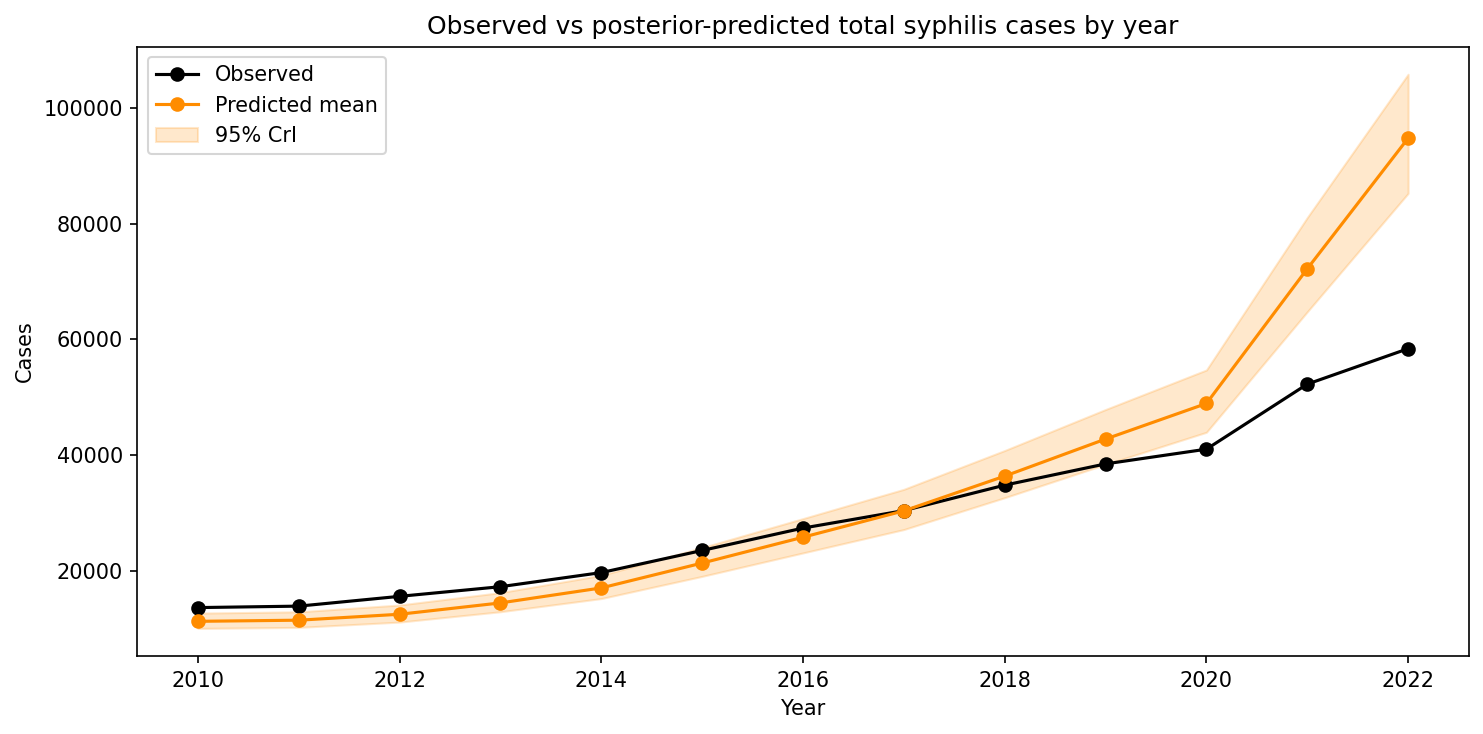

Figure S4 Yearly aggregate posterior predictive check for the main 2010–2022 model

Table S3 Comparison of observed yearly syphilis totals, yearly totals implied by the fitted mean structure (μ), and yearly totals from posterior predictive draws (y_rep)for the main 2010–2022 model

| Year | observed_total | fitted_mu_total_mean | ppc_total_mean | mu_ratio | ppc_ratio |
| --- | --- | --- | --- | --- | --- |
| 2010 | 13627 | 11253.679148803558 | 11252.04075 | 0.8258368789024406 | 0.8257166470976738 |
| 2011 | 13878 | 11441.070384171062 | 11449.00913 | 0.8244033999258583 | 0.8249754377431907 |
| 2012 | 15571 | 12480.223616122548 | 12474.37175 | 0.8015043103283378 | 0.801128492068589 |
| 2013 | 17238 | 14439.55744508876 | 14433.48888 | 0.8376585128836732 | 0.8373064668174962 |
| 2014 | 19677 | 17041.619257359693 | 17026.91863 | 0.8660679604289115 | 0.8653208631905269 |
| 2015 | 23499 | 21313.647354005134 | 21322.7895 | 0.9070023130348157 | 0.9073913570790246 |
| 2016 | 27368 | 25797.879566794363 | 25790.04938 | 0.9426293323149065 | 0.9423432247515345 |
| 2017 | 30388 | 30332.152209887885 | 30343.92613 | 0.9981621761842795 | 0.9985496289653811 |
| 2018 | 34788 | 36369.91711540573 | 36348.7935 | 1.0454730687422598 | 1.0448658589168678 |
| 2019 | 38462 | 42762.94629906671 | 42753.83413 | 1.1118232618965918 | 1.1115863482138215 |
| 2020 | 41004 | 48930.498959165256 | 48899.7625 | 1.193310383356874 | 1.1925607867525119 |
| 2021 | 52246 | 72179.7569390343 | 72182.274 | 1.3815365183752688 | 1.3815846954790798 |
| 2022 | 58373 | 94714.79901705068 | 94746.7865 | 1.622578915201389 | 1.6231268994226784 |

This temporal mismatch is likely related to the additive log-linear specification of the model, in which spatial and temporal effects enter without a space-time interaction term. The additional yearly μ-versus-PPC diagnostic showed that the discrepancy was already present in the fitted mean structure, indicating that the common year effect did not fully accommodate heterogeneous county-level temporal trajectories. Because the same year effect is applied across all counties, the model may represent rapidly increasing counties better than counties with flatter or slower growth patterns, contributing to overestimation of aggregate totals in later years.

Importantly, this aggregate-level temporal mismatch does not materially affect county-level surveillance-adjusted residual risk rankings or high-priority county classifications, which are driven primarily by the time-invariant county spatial effects rather than the shared year effects. The temporal construct validation in held-out data (HIV IRR = 3.08; gonorrhea IRR = 2.15) further supports the robustness of these county-level spatial rankings.

### 10.Leave-one-covariate-out Sensitivity Analysis(covariate)

**What it tests:** To evaluate whether each county-level covariate made a meaningful contribution to model adjustment, we re-estimated the main model five times, each time omitting one covariate while holding all other likelihood, prior, spatial, and sampling settings unchanged. Each reduced model was then compared with the main model using ΔLOOIC, the Spearman correlation of county-level residual relative risk, the Spearman correlation of posterior exceedance probability, the Jaccard overlap of the high-risk county classification, and the number of counties whose high-risk classification changed relative to the main model. This analysis therefore assesses whether the included covariates materially contributed to correction of the observed spatial pattern and to stabilization of the resulting residual risk surface.

**Interpretation:** No universal formal cutoffs are available for this sensitivity analysis, so the reduced models were interpreted relative to the main model rather than against fixed pass–fail thresholds. Here, ΔLOOIC reflects the change in approximate out-of-sample predictive fit, the Spearman correlations reflect the degree to which county-level residual risk rankings were preserved, and the Jaccard overlap reflects the extent to which the resulting high-risk county classifications matched those of the main model(16, 18, 19). Greater departure from the main model—reflected by larger ΔLOOIC values, lower Spearman correlations, and lower Jaccard overlap—indicates that the omitted covariate had made a stronger contribution to adjustment of the observed spatial pattern in the full model. Conversely, minimal change after omission suggests that the corresponding covariate contributed less to the final residual risk surface.

**Current result:** The leave-one-covariate-out results showed a clear gradient in covariate contribution (see Table S4). Omitting the social vulnerability index produced by far the largest deterioration in model fit (ΔLOOIC = +112.4), the lowest Jaccard overlap with the main model (0.694), the lowest Spearman correlation of residual relative risk (0.915), and the greatest number of changed high-risk county classifications (323 counties), indicating that social vulnerability made the largest contribution to adjustment of the observed spatial pattern. Omitting primary care physician density also worsened fit (ΔLOOIC = +17.3) and altered the high-risk classification in 157 counties, although the residual ranking remained fairly similar to the main model. By contrast, omission of insurance coverage, HPSA-derived healthcare capacity, or FQHC site density produced only limited changes in both fit and county-level classification. Together, these findings indicate that the included covariates were not redundant: social vulnerability and primary care physician density made the strongest contributions to model adjustment, whereas the remaining covariates provided more modest refinement of the resulting residual risk surface.

Table S4 Leave-one-covariate-out sensitivity analysis relative to the main model

| **Omitted covariate** | **Stably high-risk counties** | **Counties with changed high-risk classification** | **Jaccard overlap with main model** | **Spearman correlation of residual RR with main model** | **Spearman correlation of exceedance probability with main model** | **ΔLOOIC vs main model** |
| --- | --- | --- | --- | --- | --- | --- |
| Social vulnerability index | 935 | 323 | 0.694 | 0.915 | 0.901 | +112.4 |
| Primary care physician density | 855 | 157 | 0.831 | 0.974 | 0.970 | +17.3 |
| Insurance coverage | 847 | 31 | 0.964 | 0.999 | 0.998 | -3.2 |
| HPSA-derived healthcare capacity | 841 | 75 | 0.915 | 0.994 | 0.992 | +0.8 |
| FQHC site density | 857 | 55 | 0.938 | 0.996 | 0.996 | -1.8 |

### 11.Prior Sensitivity Analysis

**What it tests:** Prior sensitivity analysis evaluates whether the main conclusions are materially driven by the chosen prior specification rather than by the observed data(20). In the present analysis, the main model was refit under two alternative prior scenarios while holding the data processing, study years, county set, covariates, spatial adjacency structure, and MCMC tuning settings fixed. One scenario used wider weakly informative priors for the regression coefficients, spatial scale parameters, year effects, and overdispersion parameter, whereas the other used moderately stronger regularizing priors centered around the same general prior scale. The objective was to assess whether the county-level residual risk surface, hotspot probabilities, stably high-risk county classifications, fixed-effect incidence rate ratios, and posterior summaries of the spatial scale parameters remained stable under reasonable prior perturbations.

**Classification thresholds:** There is no single universal numeric cutoff for judging prior sensitivity, because the interpretation depends on the inferential target. In this study, meaningful prior sensitivity would have been suggested by substantial reordering of county-level residual risk rankings, marked changes in the posterior probabilities that county-level residual risk exceeded 1, low overlap between the stably high-risk counties identified under alternative priors and those identified in the main model, or notable shifts in fixed-effect incidence rate ratios and posterior spatial scale parameters. Conversely, Spearman rank correlations close to 1.0, Jaccard overlap close to 1.0, very small numbers of changed county classifications, and near-identical fixed-effect and spatial variance summaries indicate that the conclusions are robust to the tested prior choices. Because the main model and the prior sensitivity models all produced PSIS-LOO Pareto-k warnings, LOOIC differences were treated as auxiliary evidence rather than the primary basis for judging robustness.

**Current result:** To evaluate sensitivity to prior specification, the main model was refit under two alternative prior scenarios while holding the data, study years, county set, covariates, spatial adjacency structure, and MCMC tuning settings fixed. One scenario used wider weakly informative priors, whereas the other used moderately stronger regularizing priors. The alternative prior specifications used in this sensitivity analysis are summarized in Table S 5.

Table S 5 Prior sensitivity scenarios compared with the main model

| **Scenario** | **Intercept prior SD** | **Coefficient prior SD** | **Spatial scale prior SD** | **Year prior SD** | **Overdispersion prior SD** |
| --- | --- | --- | --- | --- | --- |
| Wider priors | 5.0 | 2.0 | 1.0 | 1.0 | 5.0 |
| Stronger regularization | 1.5 | 0.75 | 0.35 | 0.35 | 1.5 |

The first summary target was the stability of the stably high-risk county classification, defined as counties with residual risk ≥ 1.50 and hotspot probability ≥ 0.80. The total number of stably high-risk counties remained nearly unchanged across prior scenarios, and the overlap with the main model was extremely high (see Table S6 for more details).

Table S 6 Stability of stably high-risk county classification under prior perturbations.

| **Scenario** | **Stably high-risk counties** | **Overlap with main** | **Union with main** | **Jaccard vs main** | **Changed classifications** |
| --- | --- | --- | --- | --- | --- |
| Wider priors | 849 | 847 | 852 | 0.994 | 5 |
| Stronger regularization | 850 | 849 | 851 | 0.998 | 2 |

At the county level, both the residual risk ranking and the ranking of posterior probabilities that county-level residual risk exceeded 1 were essentially unchanged. The corresponding Spearman correlations were all extremely close to 1.0, indicating that the prior perturbations did not materially reorder counties by residual risk or posterior support for elevated risk (see Table S7).

Table S7 Stability of county-level residual risk and hotspot probability rankings.

| **Scenario** | **Spearman residual risk vs main** | **Spearman hotspot probability vs main** |
| --- | --- | --- |
| Wider priors | 0.999958 | 0.999606 |
| Stronger regularization | 0.999967 | 0.999516 |

The few changed county classifications were all threshold-adjacent cases. In every instance, the county remained very close to one of the two pre-specified cutpoints, either residual risk = 1.50 or hotspot probability = 0.80. Thus, the classification changes did not reflect broad movement in the spatial risk surface, but rather local threshold sensitivity around boundary cases (see Table S8).

Table S8 Counties with changed stably high-risk status under prior sensitivity scenarios.

| **Scenario** | **FIPS** | **County** | **Scenario residual risk** | **Main residual risk** | **Scenario hotspot probability** | **Main hotspot probability** | **Scenario stably high-risk flag** | **Main stably high-risk flag** | **Interpretation** |
| --- | --- | --- | --- | --- | --- | --- | --- | --- | --- |
| Wider priors | 06041 | Marin County | 1.504 | 1.499 | 0.994 | 0.993 | 1 | 0 | Residual risk crosses 1.50 under wider priors |
| Wider priors | 30103 | Treasure County | 2.279 | 2.279 | 0.797 | 0.805 | 0 | 1 | Hotspot probability moves just below 0.80 |
| Wider priors | 34031 | Passaic County | 1.504 | 1.500 | 0.997 | 0.998 | 1 | 0 | Residual risk crosses 1.50 under wider priors |
| Wider priors | 37185 | Warren County | 1.498 | 1.502 | 0.940 | 0.944 | 0 | 1 | Residual risk moves just below 1.50 |
| Wider priors | 53021 | Franklin County | 1.496 | 1.502 | 0.988 | 0.988 | 0 | 1 | Residual risk moves just below 1.50 |
| Stronger regularization | 06041 | Marin County | 1.503 | 1.499 | 0.992 | 0.993 | 1 | 0 | Residual risk crosses 1.50 under stronger regularization |
| Stronger regularization | 53021 | Franklin County | 1.496 | 1.502 | 0.989 | 0.988 | 0 | 1 | Residual risk moves just below 1.50 |

Posterior summaries of the spatial scale parameters were also highly stable. Under both alternative prior settings, the structured spatial component remained close to the main model estimate, and the unstructured component remained very small. This pattern supports the interpretation that the residual spatial signal was predominantly structured and was not materially driven by the tested prior choices (see Table S9).

Table S9 Posterior summaries of spatial scale parameters under prior sensitivity scenarios.

| **Scenario** | **Median sigma_structured** | **95% CrI for sigma_structured** | **Median sigma_unstructured** | **95% CrI for sigma_unstructured** | **Mean alpha_nb** |
| --- | --- | --- | --- | --- | --- |
| Wider priors | 1.459 | [1.407, 1.511] | 0.023 | [0.001, 0.072] | 4.008 |
| Stronger regularization | 1.451 | [1.400, 1.503] | 0.025 | [0.001, 0.077] | 3.998 |

The fixed-effect results were likewise nearly unchanged. Across both prior sensitivity scenarios, the incidence rate ratios for the covariates were almost identical to those of the main model, with only trivial absolute shifts. The directions of association and the overall substantive interpretation of the covariates were unchanged (see Table S10).

Table S10 Fixed-effect incidence rate ratios under prior sensitivity scenarios compared with the main model.

| **Covariate** | **Main IRR** | **Wider-prior IRR** | **Delta vs main** | **Stronger-regularization IRR** | **Delta vs main** |
| --- | --- | --- | --- | --- | --- |
| vulnerability_index | 1.5961 | 1.5962 | +0.0001 | 1.5964 | +0.0003 |
| pc_md_per100k | 1.2265 | 1.2266 | +0.0001 | 1.2261 | -0.0004 |
| insured_rate | 1.0431 | 1.0412 | -0.0019 | 1.0431 | +0.0000 |
| hpsa_capacity | 0.9098 | 0.9094 | -0.0004 | 0.9101 | +0.0003 |
| fqhc_sites_per100k | 0.9014 | 0.9005 | -0.0009 | 0.9011 | -0.0002 |

Finally, model-level PSIS-LOO summaries were compared. LOOIC values were very similar across the main and sensitivity models, but these differences should be interpreted cautiously because PSIS-LOO Pareto-k warnings were present in all three specifications. Accordingly, LOOIC was treated as a secondary diagnostic, whereas the much stronger evidence for robustness came from the near-perfect ranking correlations, the high Jaccard overlap, the minimal number of changed county classifications, and the closely matching posterior summaries for the spatial scale parameters (see Table S11).

Table S11 LOOIC and Pareto-k diagnostics for the main and prior sensitivity models

| **Model/scenario** | **LOOIC** | **Delta LOOIC vs main** | **Pareto-k > 0.7** | **Pareto-k > 1.0** | **Max Pareto-k** | **Warning** |
| --- | --- | --- | --- | --- | --- | --- |
| Main model | 117,188.219 | reference | 44 | 12 | 1.549 | True |
| Wider priors | 117,193.520 | +5.301 | 44 | 8 | 1.620 | True |
| Stronger regularization | 117,187.719 | -0.500 | 43 | 9 | 1.608 | True |

The ranking stability results are also reflected visually in the main-model versus sensitivity-model scatter plots. In both prior sensitivity scenarios, county-level residual risk estimates from the alternative model were plotted against the corresponding estimates from the main model. If the prior perturbations had materially altered the county-level ordering, substantial dispersion away from the identity line would have been expected. Instead, the points in both scatter plots clustered tightly along the y = x line, providing a visual confirmation that the county-level residual risk surface was essentially unchanged under the tested prior specifications(see fig S1and S2).

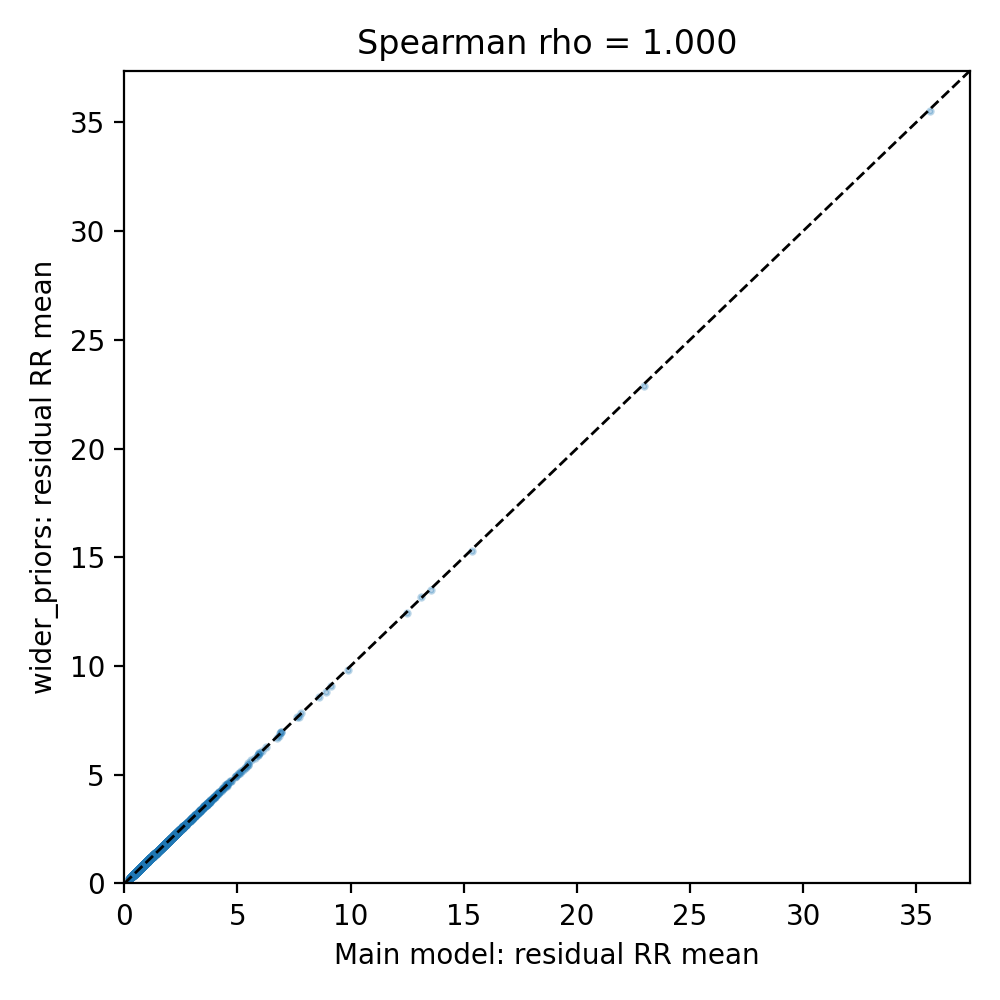

Fig 1. Main model vs wider priors residual risk

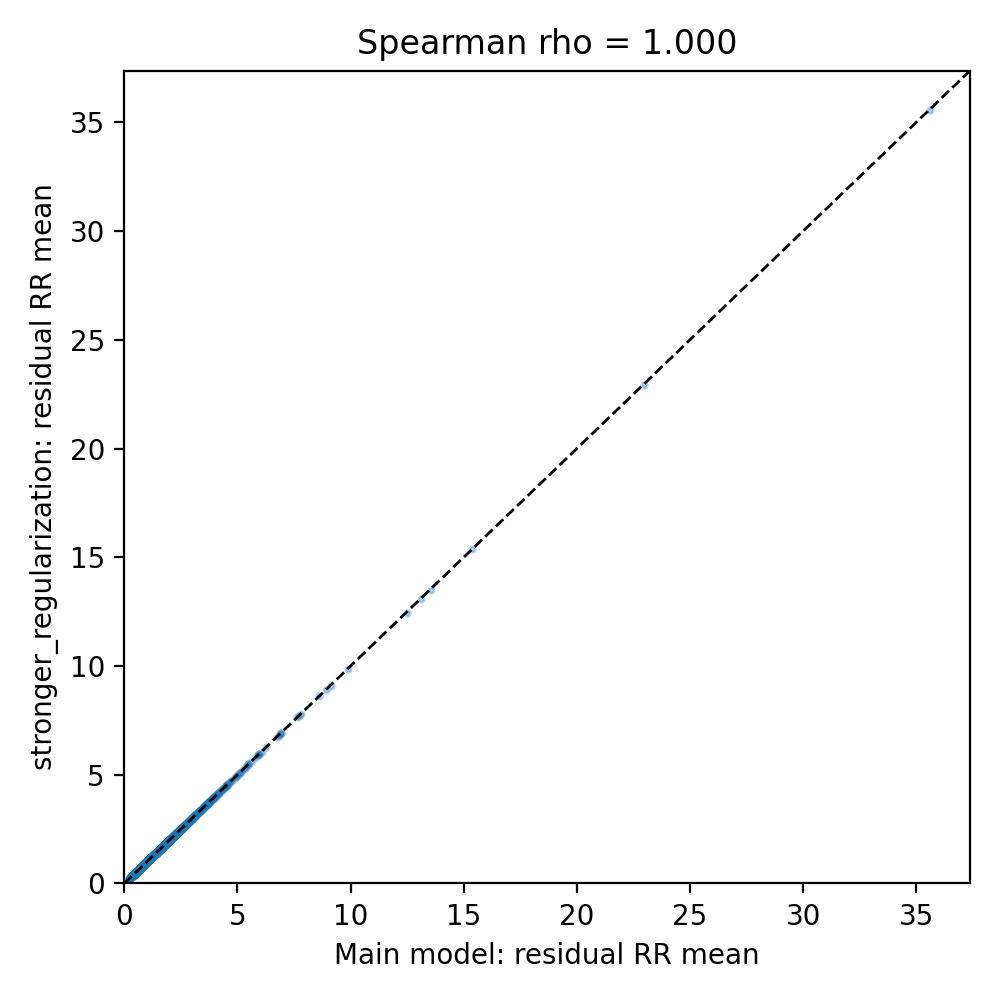

Fig 2. Main model vs stronger regularization residual risk

Overall, the prior sensitivity analysis provided strong support for the robustness of the main model. The tested prior changes did not materially affect the number or identity of stably high-risk counties, the county-level residual risk ranking, the county-level hotspot probability ranking, the fixed-effect incidence rate ratios, or the posterior summaries of the spatial scale parameters. The small number of changed county classifications occurred only among threshold-adjacent counties and therefore does not weaken the overall substantive conclusions of the study.

### 12.Summary section

Table S5 summarizes the main model diagnostic results for the primary 2010–2022 analysis.

Table S12 Summary table

| **Diagnostic** | **Purpose** | **Threshold** | **Our value** | **Status** |
| --- | --- | --- | --- | --- |
| Divergences | Sampler validity | 0 | 0 | Pass |
| Max tree depth | Sampling efficiency | No samples should reach max treedepth (14) | 7 | Pass |
| R-hat | Chain convergence | < 1.01 strict; < 1.05 acceptable | ≤ 1.016 | Acceptable |
| ESS (minimum) | Monte Carlo precision | > 400 recommended | 293 (sigma_unstructured) | Borderline |
| BFMI | Energy exploration | > 0.3 | 0.77–0.88 | Pass |
| Zero proportion | Zero-count calibration | Broad qualitative agreement | 53.2% vs 51.1% | Pass |
| Residual Moran’s I | Spatial autocorrelation | Two-sided p > 0.05 | I = 0.016, p = 0.134 | Pass |
| Pareto k (% > 1.0) | Observation influence | < 1% problematic | 0.03% (12/40,276) | Pass |
| Yearly PPC ratio | Temporal calibration | Approximately 1.0 across years | 0.83–1.62 | Flag |
